# Genome-wide association study provides novel insight into the genetic architecture of severe obesity

**DOI:** 10.1101/2025.02.25.25322870

**Authors:** Mohanraj Krishnan, Mohammad Yaser Anwar, Anne E Justice, Geetha Chittoor, Hung-Hsin Chen, Rashedeh Roshani, Roelof A.J Smit, Michael H Preuss, Nathalie Chami, Benjamin S Hadad, Esteban J Parra, Miguel Cruz, Qin Hui, Peter W.F Wilson, Yan V Sun, Xiaoyu Zhang, Gregorio V Linchangco, Sharon L.R Kardia, Jessica D Faul, David R Weir, Lawrence F Bielak, Heather M Highland, Kristin L Young, Baiyu Qi, Yujie Wang, Myriam Fornage, Christopher Haiman, Iona Cheng, Ulrike Peters, Charles Kooperberg, Steven Buyske, Joseph B McCormick, Susan P Fisher-Hoch, Frida Lona-Durazo, Jesus Peralta, Jamie Gomez-Zamudio, Stephen S Rich, Kendra R Ferrier, Ethan M Lange, Christopher G Gignoux, Eimear E Kenny, Genevieve L Wojcik, Kelly Cho, Michael J Gaziano, Luc Djousse, Shuwei Liu, Dhananjay Vaidya, Renée de Mutsert, Navya S Josyula, Christopher R Bauer, Wei Zhao, Ryan W Walker, Jennifer A Smith, Leslie A Lange, Mariah C Meyer, Ching-Ti Liu, Lisa R Yanek, Miryoung Lee, Laura M Raffield, Ruth J.F Loos, Penny Gordon-Larsen, Jennifer E Below, Kari E North, Mariaelisa Graff

## Abstract

Severe obesity (SevO) is a primary driver of cardiovascular diseases (CVD), cardiometabolic diseases (CMD) and several cancers, with a disproportionate impact on marginalized populations. SevO is an understudied global health disease, limiting knowledge about its mechanisms and impacts. In genome-wide association study (GWAS) meta-analyses of the tail end of the BMI distribution (≥95^th^ percentile BMI) and two SevO phenotypes [Obesity Class III BMI ≥40 kg/m^2^ and Obesity Class IV BMI ≥50 kg/m^2^] in 159,359 individuals across eleven ancestrally diverse population-based studies followed by replication in 480,897 individuals across six ancestrally diverse studies, we identified and replicated one novel signal in an unknown locus [*BHLHE40-AS1*] and three novel signals in known loci of BMI [*TENM2*, *PLCL2*, *ZNF184*], associated with SevO traits. We confirmed a large overlap in the genetic architecture of continuous BMI and severe obesity phenotypes, suggesting little genetic heterogeneity in common variants, between obesity subgroups. Systematic analyses combining functional mapping, polygenic risk scores (PRS), phenome wide association studies (PheWAS) and environmental risk factors further reinforce shared downstream comorbidities associated with continuous measures of BMI and the importance of known lifestyle factors in interaction with genetic predisposition to SevO. Our study expands the number of SevO signals, demonstrates a strong overlap in the genetic architecture of SevO and BMI and reveals a remarkable impact of SevO on the clinical phenome, affording new opportunities for clinical prevention and mechanistic insights.

## MAIN

Severe obesity (SevO) (Body Mass Index [BMI] (≥40 kg/m^2^) is an emerging global health disease which imparts a substantial and growing morbidity and mortality burden that disproportionately affects underserved populations^1–3^. With marked heterogeneity in the disease of obesity, severe obesity represents a higher degree of obesity than frank obesity, more extreme weight gain, and potential mechanistic differences ^4^. Although SevO has been associated with a myriad of health complications including type 2 diabetes (T2D), coronary heart disease (CHD) and various forms of cancer^5,6^, very few studies have explored the biological and genetic mechanisms underlying SevO. Indeed, most studies have grouped individuals with SevO with individuals classified according to World Health Organization (WHO) class I ([BMI] ≥30 kg/m^2^ to <35 kg/m^2^) or class II ([BMI] ≥35 kg/m^2^ to <40 kg/m^2^) obesity, or they have been excluded altogether in clinical and epidemiological studies^7^, thereby masking possible detrimental effects of the critical SevO subtype. Thus, the health impacts for individuals at the highest end of the spectrum for obesity are underestimated or largely undocumented, limiting knowledge about the mechanistic pathways and impacts, including its genetic determinants. Genome wide association studies (GWAS) have mapped thousands of independent loci among people of diverse ancestries influencing both dichotomous and quantitative traits of BMI ([BMI] ≥30 kg/m^2^)^8–12^, offering insights into the genetic architecture of obesity. To date, the largest known population-based GWAS of SevO was performed in 263,407 individuals of European ancestry and identified seven novel loci associated with different classes of obesity; two of which (*HS6ST3* [heparan sulfate 6-O-sulfotransferase 3] and *ZZZ3* (zinc finger ZZ-type containing 3) were associated with WHO Obesity Class II ([BMI] ≥35 kg/m^2^)^13^. The remaining loci associated with BMI tails and clinical classes of SevO had variants that intersected with studies of continuous BMI GWAS^12,14^ and mapped highly penetrant variants in loci that affect key molecular and neural pathways involved in human energy homeostasis^13,15–22^. Given that most SevO GWAS have been limited by small sample sizes^16,23^, inconsistent phenotype definition (clinical classes of obesity, extremes of distribution tails)^24^, a lack of participants of diverse backgrounds (with current analyses mainly including Europeans)^23,25^, and a focus on monogenic causes of obesity that are difficult to detect in population-based samples,^13^ we sought to perform the largest known GWAS of SevO among individuals of diverse ancestries, leveraging extremes of obesity and WHO described clinical obesity classes. We hypothesized that given its extreme form, common variants for SevO could be mapped with smaller sample sizes than GWAS for other obesity related traits and that such variants would likely overlap with variants mapped using continuous BMI and other obesity binary traits. Lastly, we performed functional mapping, polygenic risk score analysis (PRS), phenome wide association studies (PheWAS), and assessed environmental risk factors in the context of predicted high risk for, to discern the biological mechanisms underlying body weight regulation.

## RESULTS

### Demographic characteristics

Genotyping, imputation quality control, and statistical platforms used for analyses are detailed in **Supplementary Table 1**. We defined cases as individuals in the top 5th percentile of BMI, and controls in the 5th–50th percentile, stratified by sex after adjusting for age, age², and ancestry (BMI cases ranged from 34 kg/m^2^ to 48 kg/m^2^ and BMI controls ranged from 22 kg/m^2^ to 27 kg/m^2^) (**Supplementary Figures 1-4**). We also used the WHO classification to define SevO cases as individuals with BMI ≥40 kg/m² (Obesity Class III) or ≥50 kg/m² (Obesity Class IV), and controls as a BMI of 18.5–24.9 kg/m². We defined loci as index SNPs meeting genome-wide significance (P ≤5×10⁻D) and SNPs within ±250kb in linkage disequilibrium (LD, r² <0.6) using the 1000 Genomes Project reference. If multiple independent SNPs (r² <0.1) reached significance within a locus, each was functionally annotated. We also considered rare variants (MAF <1%) given their potential higher frequencies in ancestry-stratified analyses, though acknowledging the unreliability of rare variants genotyped by SNP arrays. Stage 1 discovery and Stage 2 replication analyses included 11 and 6 population-based studies, with a total of 159,359 and 480,997 individuals across diverse ancestries (**Figure 1; Supplementary Tables 2and 3**).

**Figure 1:**
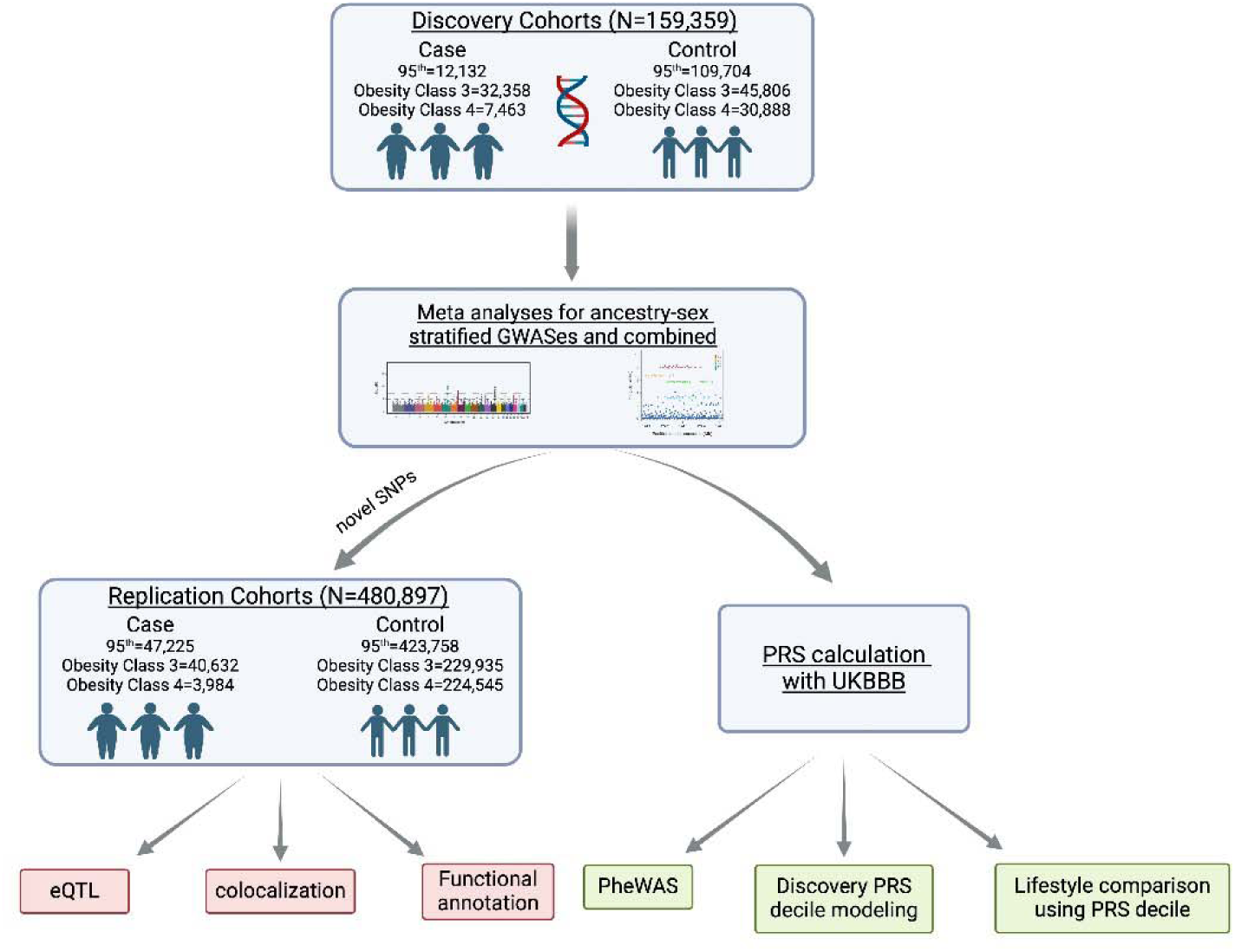
Schematic of the study design for genomic analysis and systematic comparisons of SevO in ancestrally diverse populations.

### GWAS of severe obesity (SevO) traits

We analyzed >31 million genotyped or imputed SNPs in ancestry-, sex-specific, and combined analyses (**Supplementary Figures 5-13**). In the combined ancestry and sex analysis, we identified four novel signals in or near unknown loci of obesity (*BHLHE40-AS1*, *PLAS2R1*, *PIWL1* and *CDHR1*) and six novel signals in or near known obesity loci (*PDGFC, SMARCAD1, TENM2, ZNF184, PLCL2,* and *SLC39A11)* at genome-wide significance (P <5×10⁻D) (**Supplementary Table 4**). SNP-based heritability in the discovery phase ranged from 0.27 to 0.29 for WHO Class III and 0.37 to 0.52 for Class IV obesity (**Supplementary Table 24**). Both SevO classes showed high genetic correlation with BMI (rg = 0.94 for Class III, 0.93 for Class IV). These ten variants were carried forward to Stage 2 replication. We replicated the novel *BHLHE40-AS1* signal and three novel signals near known loci (*TENM2, ZNF184, PLCL2*) in a meta-analysis using a Bonferroni-adjusted threshold of *P* < 0.005 (**Table 1**; **Figure 2**). All variants showed consistent effect directions across SevO classes. Ancestry-stratified summary statistics for these 10 variants are in **Supplementary Tables 5-10**.

**Table 1:** Summary of unreported independent GWAS signals of severe obesity traits in the discovery and replication cohorts. Independent signals were defined as lead variants having met a genome-wide significant *P* value (*P*□<□5□×□10^−8^) in our discovery cohort and a Bonferroni adjusted significance (*P=*0.005) in the replication cohort across any strata (all, men, or women) had a linkage disequilibrium (LD) *r*^2^□<□0.6 with other variants and were defined as those independent from each other at *r* ^2^ < 0.1 inside a subset of independent significant variants. Genes were annotated based on ±250kb of the lead variant and/ or closest gene present.

| Marker (Chr:Position hg19:<br>Effect Allele:<br>Other Allele) | Genes<br>($\pm 250\text{kb}$ of<br>the lead<br>variant) | rsid | *Discovery<br>95% tile | OR | †Replication<br>95% tile | OR | *Discovery<br>ObClass3 | OR | †Replication<br>ObClass3 | OR | *Discovery<br>ObClass4 | OR | †Replication<br>ObClass4 | OR |
| --- | --- | --- | --- | --- | --- | --- | --- | --- | --- | --- | --- | --- | --- | --- |
| 3: 5944942:T:C | BHLHE40-AS1 | rs17824177 |  |  | All, Women,<br>Men | ↓ | All | ↓ | All, Men | ↓ |  |  |  |  |
| 3: 17115122:C:G | PLCL2 | rs36118680 |  |  | All | ↑ | All | ↑ | All | ↑ |  |  |  |  |
| 5: 166756351:T:C | TENM2 | rs13155681 | All | ↑ | All, Women | ↑ |  |  | All, Men | ↑ |  |  |  |  |
| 6: 27449233:A:G | ZNF184,<br>ZNF391,<br>POM121L2<br>PRSS16 | rs140919115 |  |  | All, Men | ↑ | Women | ↑ | All, Men | ↑ |  |  | Women | ↑ |

\* Discovery Significance is determined by GWAS significance threshold ( $5 \times 10^{-8}$ )
Note initial GWAS using the discovery cohort identified 10 novel variants associated with severe obesity classes. Here we show the 4 variants that were identified in our discovery cohort and were validated in our replication cohort using Bonferroni adjusted threshold of 0.005. For full results for all 10 variants for both discovery and replication cohort please ST4. The arrows denote relative odds of the occurrence of the outcome of interest using 1 as the reference.

**Figure 2:**
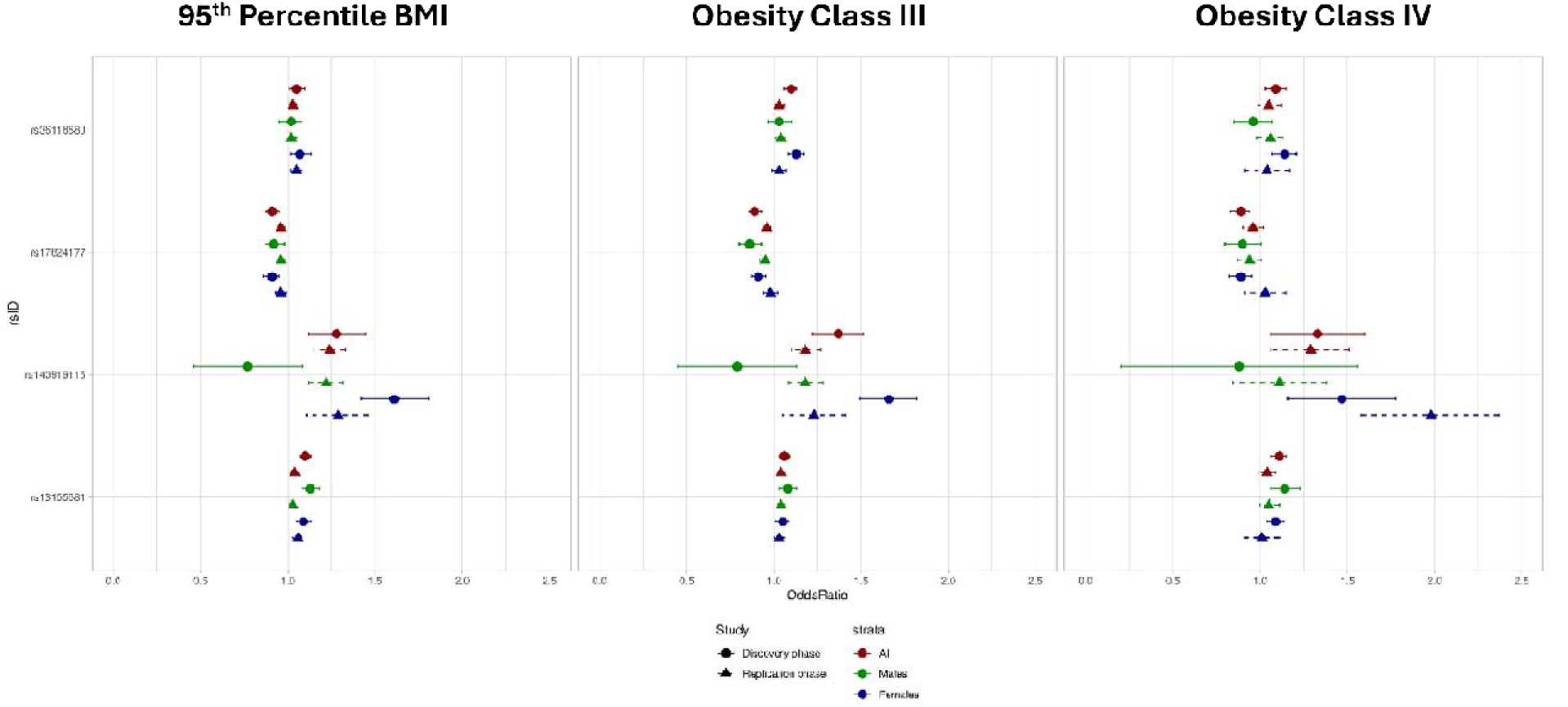
Forest-plot illustrating the direction of association for the four validated variants (discovery and replication) across the three obesity classes (95% class, Obesity Class 3 and Obesity Class 4) for each stratum (all, females and males).

### Expression Quantitative Trait Loci (eQTL), colocalization and functional annotations of novel GWAS findings

We evaluated the functional and clinical significance of our four variants identified in Stage 1 discovery and validated in Stage 2 replication using colocalization with expression quantitative trait loci (eQTLs) from GTEx^26^, eQTLGen Phase 1^27^, and a large Mexican American whole blood eQTL resource. No eQTLs were found in eQTLGen Phase 1. In GTEx, the minor (C) allele of rs36118680 was associated with increased *PLCL2* expression in whole blood (NES = 0.28, *P* = 6.3×10⁻¹D) (**Supplementary Table 11a**). In the Mexican American dataset, the minor (C) allele of rs17824177 was associated with higher *BHLHE40-AS1* expression (NES = 0.395, P = 0.01) **(Supplementary Table 11b).** Colocalization modeling with Obesity Class III GWAS data revealed colocalization between rs36118680 and *PLCL2* expression in whole blood (posterior probability = 99%).

### Plausible biological roles for genes surrounding novel replicated variants

Many of the genes surrounding these index variants (±250kb of the index variant or closest gene present) had important roles in the pathogenesis of SevO including neuronic control of food intake, sustained low-grade chronic inflammation and adipogenesis. A summary of the plausible biological roles of genes surrounding the four novel variants with SevO using online resources and a comprehensive manual review of the published literature is presented in **Supplementary Table 12.**

### Overlap with known BMI loci

We assessed the relevance of known BMI loci for Obesity Class III by examining 813 established BMI-associated variants (*P* <5×10⁻D) in our ancestry-combined Obesity Class III discovery results. Of these, 464 variants (57.07%) showed consistent direction and nominal significance (P <0.05) with Obesity Class III, indicating a strong enrichment of susceptibility alleles across traits (R Pearson = 0.89, P = 2.02×10⁻²DD) (**Supplementary Figure 15**, **Table 17**). We also analyzed our four validated novel variants (or their LD proxies, R² >0.8) in the GIANT+UKBB^12^ and UKBB trans-ancestry BMI meta-analyses. In GIANT+UKBB by Yengo *et al*, only rs17824177 showed some evidence of association with BMI (*P* = 2.6×10⁻D), though not genome-wide significant. The other variants were absent due to their rarity or the older HapMap imputation panel. In the UKBB trans-ancestry meta-analysis, genome wide evidence of association with BMI was not demonstrated (rs17824177: *P* = 0.00112, rs36118680: *P* = 0.0118, rs13155681: *P* = 0.132, rs140919115: *P* = 0.246).

### PRS-estimation of SevO

PRS were calculated using two methods: PRS-CS with HapMap3 SNPs for SevO traits (e.g., 95^th^ percentile BMI, Obesity Class III, and Class IV) and BMI GWAS results from the GIANT consortium (N∼300k^14^), and genome-wide significant independent variants from SevO GWAS traits in this study. We tested the PRS on phenotypic and genotype data from 235,071 unrelated UKBB participants and 3,063 unrelated Mexican Americans from the CCHC study, all classified as cases or controls for SevO. The participants included 223,699 Europeans, 4,057 African or African Americans, 1,537 East Asians, 4,978 South Asians, and 3,063 Mexican Americans. PRS performance by population is shown in **Figure 3** and **Supplementary Table 15**.

**Figure 3:**
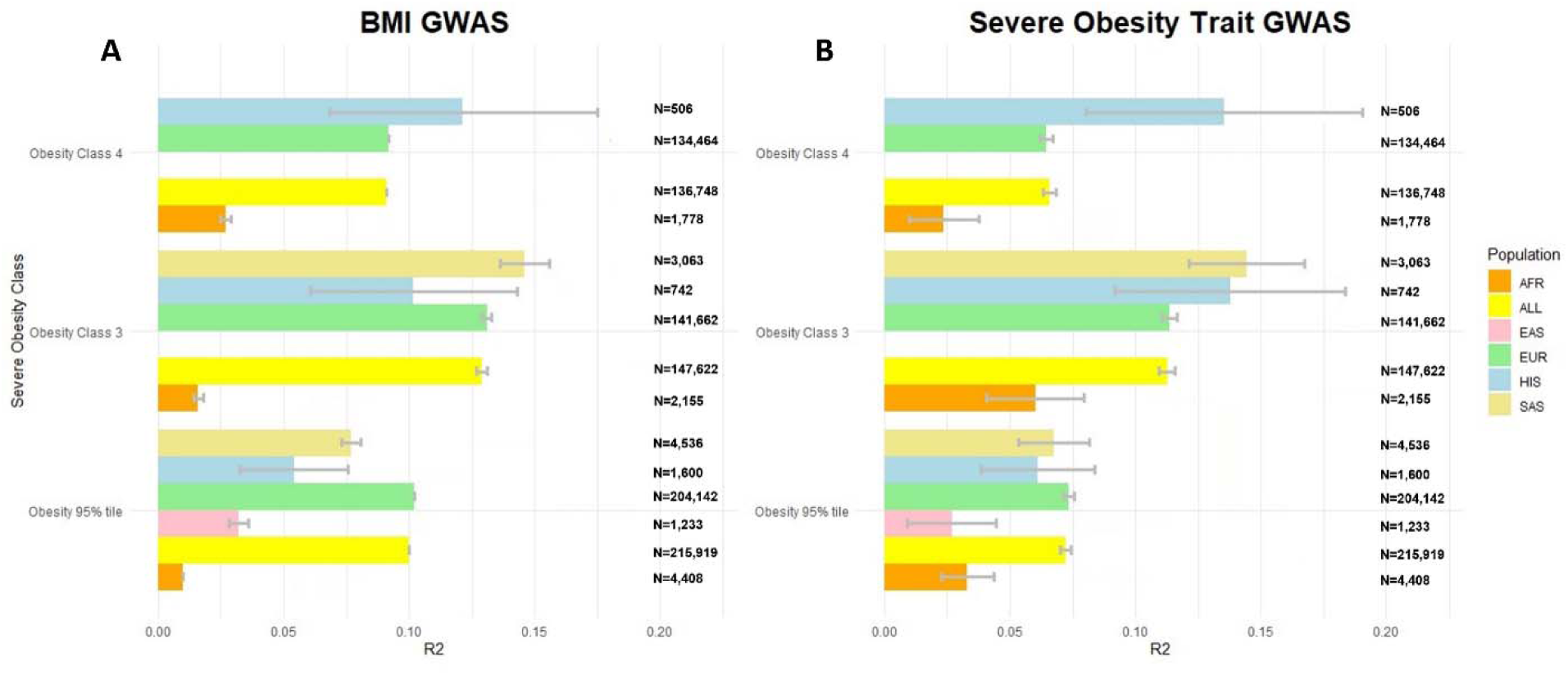
Phenotypic variance explained (R^2^) by **(A)** using HapMap 3 SNPs in the BMI GWAS from the GIAN consortium and **(B)** using the PRS-CS method with HapMap3 consortium SNPs for each of the SevO GWAS traits (e.g., 95% Obesity, Obesity Class III and Obesity Class IV). Using approximately 1/3 the sample size in our SevO PRS we show similar predictive power in explaining phenotypic variance of SevO traits when compared to the PRS generated from the BMI GWAS.

As expected, R² estimates were low when using only GWAS-significant SNPs to calculate PRS. However, the PRS-CS method with HapMap 3 variants improved R² across all ancestries. For Obesity Class III, R² improved to 6.0% for African participants, 11.4% for Europeans, 13.8% for Hispanics, and 14.4% for South Asians. For the 95th percentile BMI, R² was 3.3% for Africans, 2.7% for East Asians, 7.4% for Europeans, 6.1% for Hispanics, and 6.8% for South Asians. For Obesity Class IV, R² was 2.4% for Africans, 6.5% for Europeans, and 13.5% for Hispanics (**Supplementary Table 15**). Overall, PRS performance was highest for Obesity Class III, except in the Hispanic population. We also calculated PRS using BMI GWAS results and compared the performance to SevO GWAS-based PRS. R² performance was similar across ancestries, though the SevO GWAS had a smaller sample size (∼120,000 for the 95th percentile, ∼80,000 for Obesity Class III, ∼50,000 for Class IV) compared to the BMI GWAS (∼300,000) (**Supplementary Table 15**). Only three East Asian participants had BMI ≥40 kg/m², and no East or South Asian participants had BMI ≥50 kg/m², so PRS were not calculated for these groups.

### PRS-PheWAS in the UKBB

We conducted a phenome-wide association study (PheWAS) of PRS-SevO across diverse populations in the UKBB (**Supplementary Figure 14; Supplementary Tables 16-20**). Obesity Class III PRS was associated with 37% of phenotypes (out of 1,668) in Europeans, including known obesity-related traits like BMI, body fat, waist circumference, metabolic comorbidities, and bone density. We also found strong associations with inflammation markers (C-reactive protein, gamma-glutamyltransferase) and hematopoietic traits (immature reticulocyte fraction, reticulocyte count) (**Figure 4; Supplementary Tables 16-20**).

**Figure 4:**
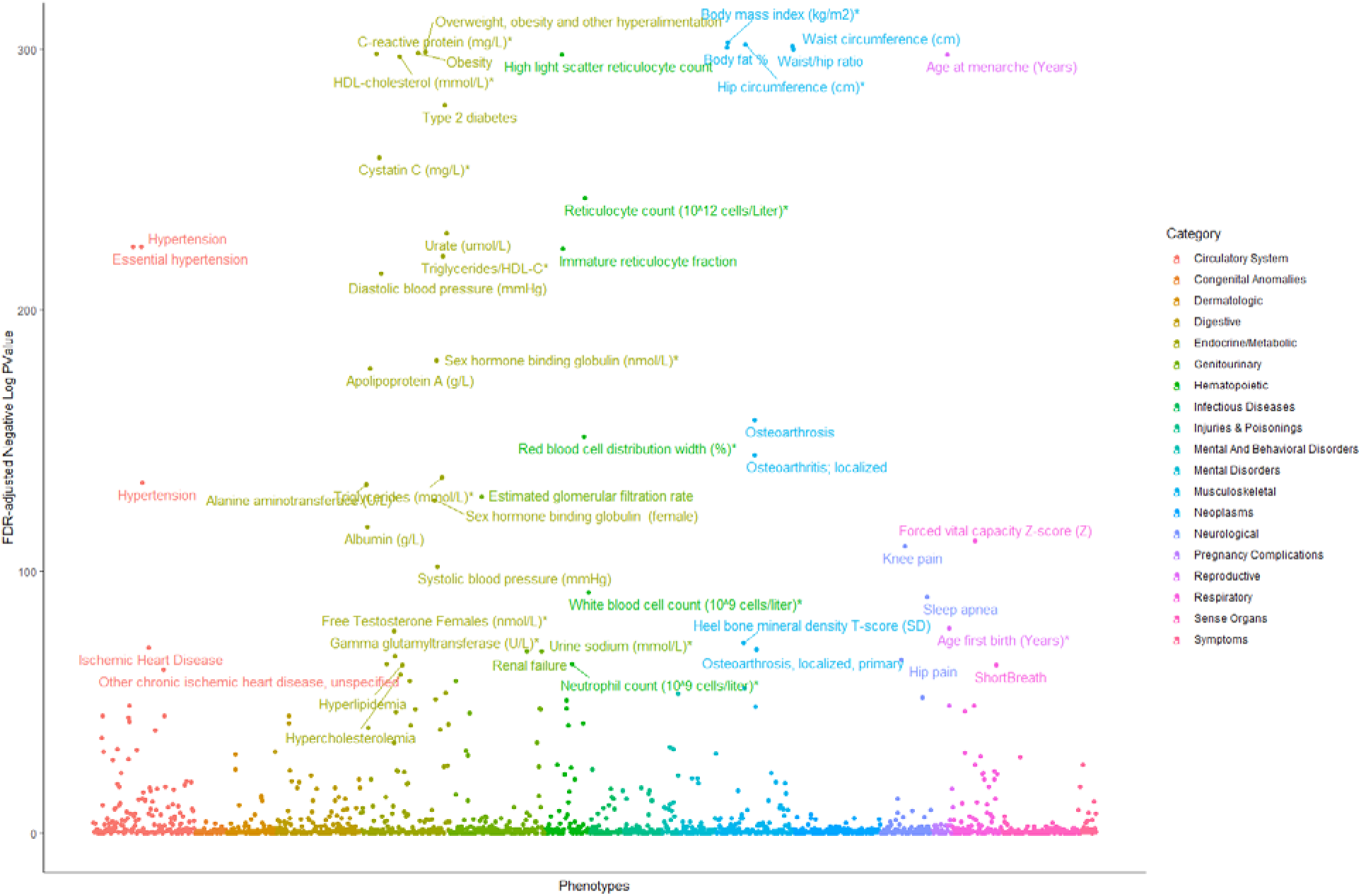
PheWAS analysis of secondary phenotypes (max 1668) within the UKBB Europeans with our Obesity Class III PRS derived from our discovery analyses.

### PRS deciles and lifestyle behavior modelling

A recent PRS for BMI, based on 2.1 million genetic variants, identified individuals at risk of SevO^28^. Khera et al. found that those in the top 10% (90th percentile) of PRS had an average BMI 2.9 kg/m² higher than those in the lower 90%. We replicated this approach with our PRS-CS, separating them into deciles (1-10) according to BMI categories (1) BMI <25 kg/m^2^, 2) BMI ≥25 kg/m^2^ & BMI <30 kg/m^2^, 3) BMI ≥30 kg/m^2^ & BMI <40 kg/m^2^ and 4) BMI ≥40 kg/m^2^) among diverse ancestries in the UKBB (**Supplementary Table 21**). We then assessed the predictive power of our SevO PRS (90^th^ percentile vs. 10^th^ percentile) on Obesity Class III, in our ancestry -specific and -combined samples from the UKBB (**Supplementary Table 22; Supplementary Figures 16 and 17**). Focusing on the combined samples, we found that SevO was present in 4.9% of those >90^th^ percentile for PRS compared to 0.55% of those in <10^th^ percentile category, corresponding to a 10-fold increased risk of SevO (**Figure 5**). These differences in distribution indicate a different pattern of BMI distribution across the tail ends of the PRS distribution. **Supplementary Table 25** provides estimates of the specificity, sensitivity, and positive and negative predictive values of our SevO PRS for Obesity Class III across the European, African, and South Asian samples from UKBB and the Hispanic samples from the CCHC cohort. The predictive power of the SevO PRS, assessed using receiver operating characteristic (ROC) curves, showed AUC values ranging from 0.62 to 0.77 when using PRS alone, and improved to 0.75 to 0.83 when combined with covariates across all ancestries. Sensitivity for Obesity Class III ranged from 15% to 39%, with positive predictive values from 16% to 33%.

**Figure 5:**
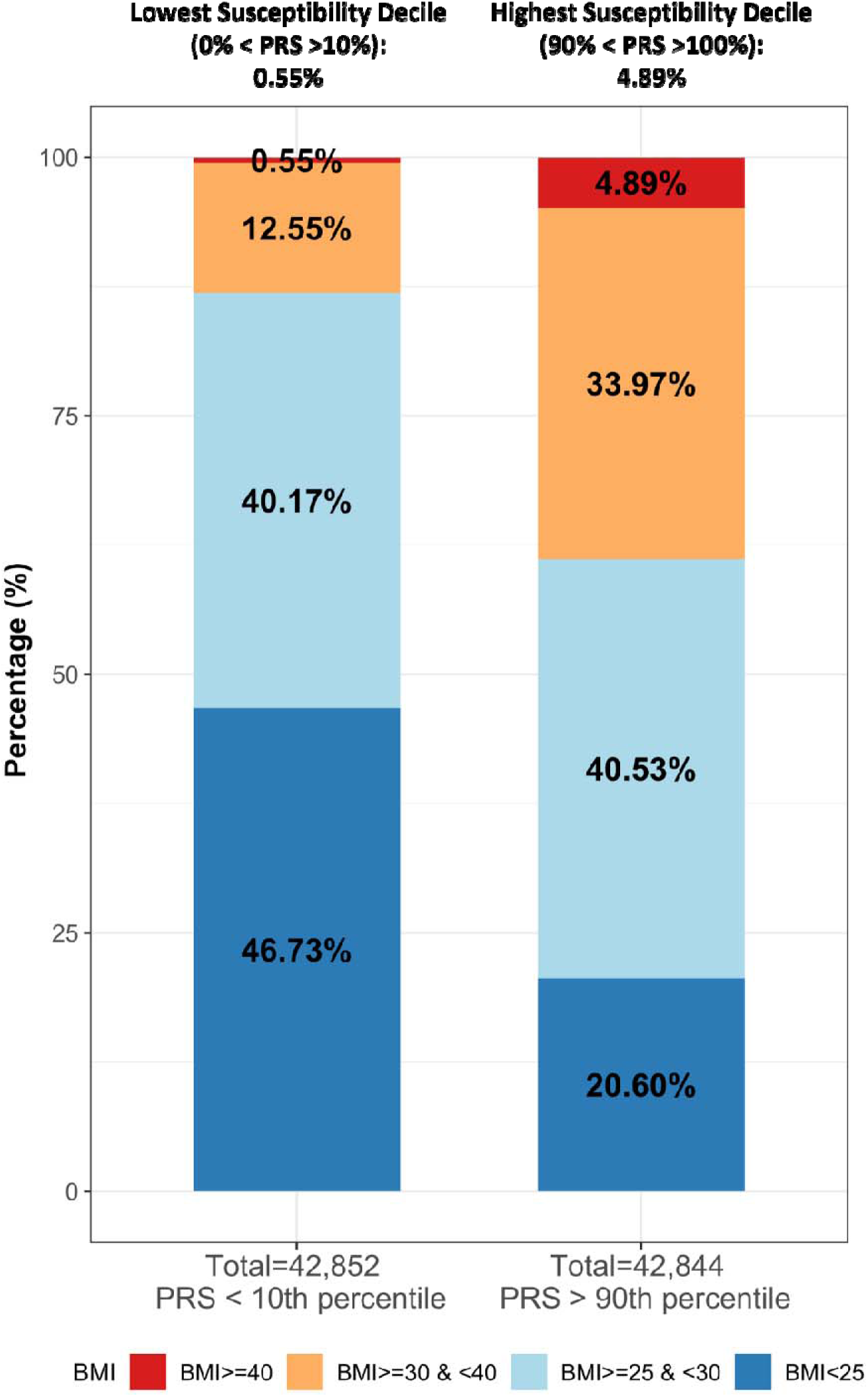
Relationship of SevO PRS > 90^th^ percentile with < 10^th^ percentile across BMI categories in ancestry - combined samples from the UKBB.

We also compared the pattern of association of individual lifestyle factors with BMI, within each of the PRS groups (**Figure 6; Supplementary Table 23**). Of particular interest were participants with a PRS in the ≥ 90^th^ percentile and a BMI <25 kg/m^2^, which we labeled the “Resilient group”. We compared this “Resilient group” to those with 1) in any PRS category and with SevO, 2) >90^th^ percentile for PRS and Obesity Class III, 3) < 10^th^ percentile for PRS and SevO 4) <10^th^ percentile and BMI <25 kg/m^2^. The “Resilient group” reported significantly healthier lifestyle behaviors when compared to participants with SevO or even those with BMI <25 kg/m^2^ and with PRS <10^th^ percentile, including improved dietary patterns (lower meat intake, increased fruit, vegetable, fiber and fish intake), increased physical activity and sleep duration and insomnia (**Figure 6; Supplementary Table 23**). The “Resilient group” also smoked slightly more and consumed more alcohol than participants with SevO but consumed less alcohol than those with BMI <25 kg/m^2^ and a PRS in the <10^th^ percentile range (**Figure 6; Supplementary Table 23**). The “Resilient group” was also less likely than participants with SevO but more likely than those with BMI <25 kg/m^2^ and a PRS in the <10^th^ percentile range to say they were ‘plumper’ than their peers at age 10 years (**Figure 6; Supplementary Table 2**3.

**Figure 6:**
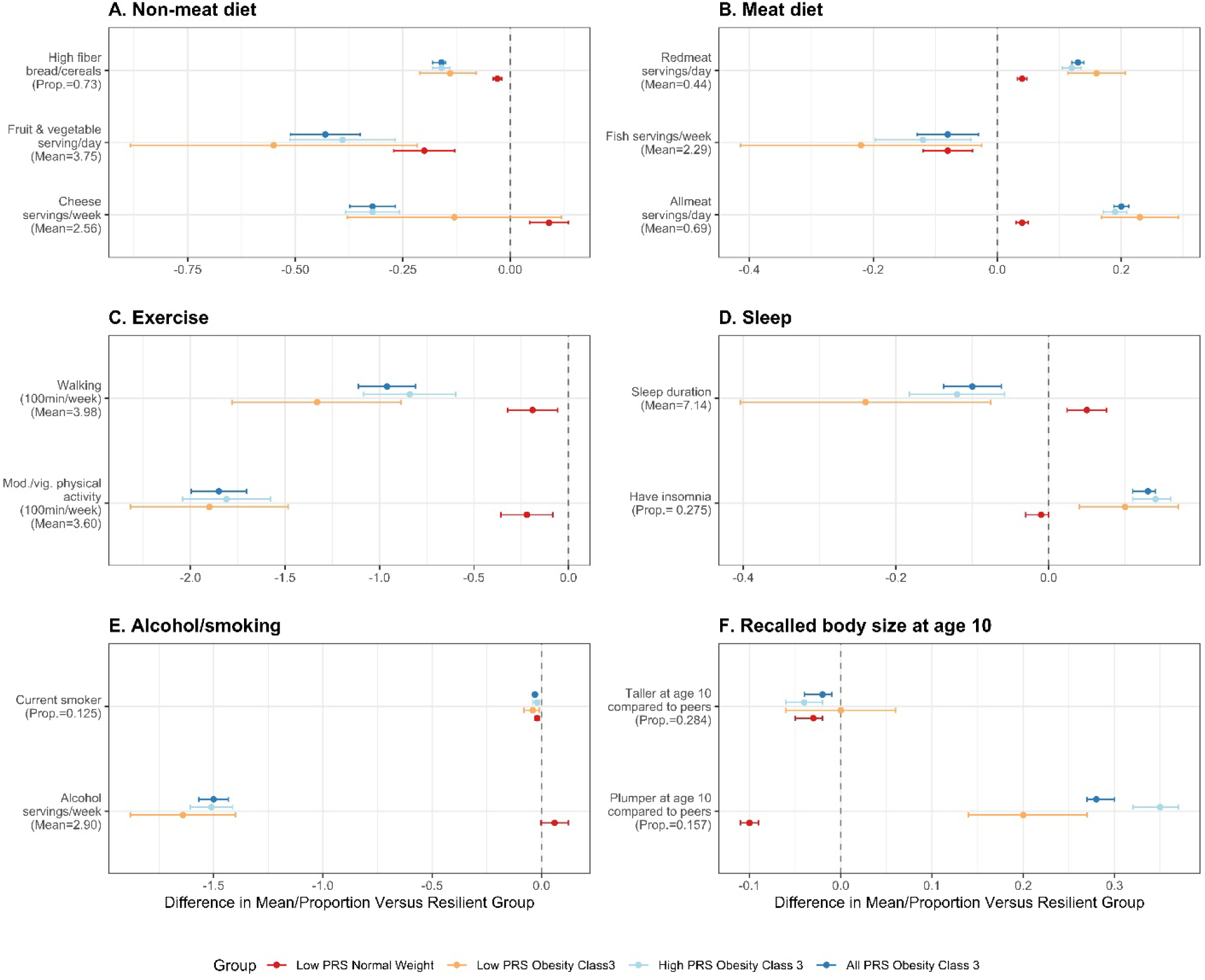
Lifestyle adherence comparisons between sample groups defined by PRS upper and lower deciles and BMI categories. The “Resilient group” was designated as individuals having a PRS in the ≥ 90^th^ percentile and a BMI <25 kg/m^2^. This group was compared to those with 1) all PRS and SevO, 2) >90^th^ percentile for PRS and SevO Class III, 3) < 10^th^ percentile for PRS and SevO and 4) <10^th^ percentile and BMI <25 kg/m^2^ with different lifestyle factors including dietary, physical activity and sleep patterns, alcohol and smoke servings and perceived body size at age 10.

## DISCUSSION

In our meta-analysis of GWAS of up to 159,359 individuals across 11 ancestrally diverse population-based studies, we identified numerous loci associated with SevO, many of which overlap with Class I obesity (BMI ≥30 kg/m^2^). Ten unreported independent variants in novel and known loci were associated with obesity related traits for the first time; four of these were validated in our replication cohort of 480,897 individuals of diverse ancestries. Of our replicated signals, rs17842177 is in a novel locus (*BHLHE40-AS1*), whereas the other three novel signals (rs36118680, rs13155681 and rs140919115) are in known BMI loci. Our study expands the number of identified SevO signals, confirms strong overlap in the genetic architecture of SevO and BMI and reveals a remarkable impact of SevO on the clinical phenome, affording new opportunities for mechanistic insights and clinical prevention and intervention. These findings further enhance the underlying biology of SevO and highlight novel genetic signals not previously implicated in SevO pathophysiology.

Before severe obesity became as common as it is today, the focus was on monogenic forms of the disease. It is only more recently that the form of common complex severe obesity became more prevalent in the population. Thus, early genetic studies of severe obesity (SevO) focused on identifying gene-disrupting mutations in the leptin-melanocortin pathway (e.g., *LEPR, MC4R, POMC*) ^29–33^, which are linked to early-onset SevO through dysregulation of food intake and food preferences^34^. While these studies provided novel mechanistic insights, GWAS have shifted focus to polygenic forms of obesity. To date, GWAS has identified over 1,000 loci influencing BMI, with some high-impact variants near genes associated with extreme and early-onset obesity ^11,14,35–38^ also enriched in polygenic obesity traits.

GWAS of SevO found numerous susceptibility loci that intersected with earlier-identified BMI loci including the archetypal *FTO, MC4R*, *SH2B1* and *NPC1* suggesting little etiological heterogeneity between obesity subgroups^16,17,19^. However, many of these GWAS had small sample-sizes, low coverage of markers tested, and inconsistent phenotype groupings, all which may have limited study findings. A large GWAS involving 263,407 individuals of European ancestry identified 165 genetic loci associated with BMI, height, waist-to-hip ratio, and WHO-defined obesity classes: Overweight (BMI ≥25 kg/m²), Obesity Class I (BMI ≥30 kg/m²), Obesity Class II (BMI ≥35 kg/m²), and Obesity Class III (BMI ≥40 kg/m²). Of these, only one known variant, rs1421085 in the *FTO* gene, was significantly associated with Obesity Class III [OR=1.47, P=2.11 × 10⁻¹D], which was identified in smaller SevO GWAS studies and replicated in our study. Additionally, 22 variants were associated with Obesity Class II, including two novel loci in *HS6ST3* (heparan sulfate 6-O-sulfotransferase 3) and ZZZ3 (zinc finger ZZ-type containing 3). The absence of novel associations with Obesity Class III in this study likely reflects low statistical power, with 3,986 cases and 67,010 controls. In our discovery GWAS, we found validation for the novel Obesity Class III traits reported by Berndt *et al.* for *HS6ST3* (rs7989336) and *ZZZ3* (rs17381664), particularly in Obesity Class IV, as well as in ancestry-combined and female-specific samples.

Consistent with findings in other polygenic traits, our study shows that large sample sizes are needed to identify variants associated with population extremes^13,39^. Using a population-based design, we identified a novel locus (*BHLHE40-AS1*) and three unreported independent signals in known BMI loci linked to SevO traits. These loci are involved in inflammation, food intake regulation, and altered adipose differentiation. Despite the strong genetic overlap between BMI and SevO (r_g_s > 0.80), focusing on trait extremes allowed us to detect novel signals with smaller sample sizes.

Obesity is often characterized by low-grade inflammation that is associated with a sequalae of metabolic diseases including type 2 diabetes, hypertension, and cardiovascular complications^40^. As an acute-phase reactant to inflammation and infection, C-reactive protein (CRP) is associated with obesity^41^ along with inflammatory mediators such as *TNF-*α and *IL-*6^42^. Our study identified the novel *BHLHE40-AS1* as a strong candidate gene of SevO both through our GWAS findings and replication but also through eQTL evidence in whole blood samples from Mexican American CCHC participants^43^. A CADD score > 10 was also reported for rs17824177 (C_score_=15.27) suggesting some level of deleteriousness (**Supplementary Table 13**). Recently, a study has demonstrated that *BHLHE40-AS1* modulates pro-inflammatory cytokines and is an important mediator of IL6/STAT3 signaling^44^. In addition, common variants in *BHLHE40-AS1* associate with gamma glutamyl transferase, a biomarker positively correlated with C-reactive protein^45^ and increased oxidative stress^46^, which is supported by our PheWAS of Obesity Class III trait in UKBB. Our study suggests that *BHLHE40-AS1* is a strong biological candidate of obesity.

Accumulating evidence also suggests that distinct psychiatric disorders including schizophrenia and major depressive disorders share a common genetic etiology with obesity^47^. The intronic variant, rs140919115 (n.265+6212G>A), located in *ZNF184* was associated with Obesity Class III. This variant surrounds a locus that harbor genes (*ZNF391*, *POM121L2* and *PRSS16I*) involved with a plethora of mood-related disorders (**see Supplementary Table 12**)^48,49^. This suggests a multi-faceted association between psychiatric disorders and BMI, involving pathways that may influence ‘binge’ eating and canonical food-related behaviors. In addition, *ZNF184* was found to amplify *FTO* gene expression levels^50^, and may regulate body through *FTO*-mediated browning and mitochondrial thermogenesis^51^.

Two additional loci (**Supplementary Table 12**) highlight genetic mechanisms involved in the developmental shift from brite (brown) adipocytes to energy-storing white adipocytes leading to reduced thermogenesis and increased lipid deposition^34^. The intronic variant, rs13155681 (c.226+44283C>T) located in *TENM2* was associated with the 95^th^ percentile BMI and may mediate obesity through adipocyte differentiation. *TENM2*, involved in regulating synaptic plasticity of neurons, is known to maintain white adipocyte phenotype, and reduced mitochondrial respiration during adipogenic differentiation^52^ leading to increased storage of fat. The intronic variant rs36118680 (c.3204+5565C>G) was associated with Obesity Class III. The *PLCL2* locus mediates several biological functions, particularly those related to protein phosphatases. and is known to modulate obesity through fat lipolysis and thermogenesis regulation of adipocytes^53,54^. However, to translate genomic findings to meaningful outcomes, incorporation of other ‘omic’ platforms combined with advanced computational tools are needed to provide more proximal insight into the dysregulation of SevO biology in response to genetic modifications.

The SevO PRS in UKBB explained 11.4%, 6.0%, 13.8% and 14.4% of the phenotypic variance for Europeans, Africans, Hispanics and South Asian participants, respectively and was similar with the PRS generated using HapMap 3 SNPs in the BMI GWAS from the GIANT consortium^14^. Notably, we had similar predictive power and performance in BMI risk assessment with just 1/3 the sample size across all ancestries. In applying our SevO PRS in PheWAS, 37% of phenotypes demonstrating significant associations (max 1668). These findings highlight a broader impact of SevO on disease morbidity and mortality than previously anticipated. Notable associations were also found with hematopoietic phenotypes, such as immature reticulocyte fraction and reticulocyte count. Genes upregulated in obesity are selectively expressed in reticulocytes^55^, aligning with studies showing higher red blood cell counts in obese individuals^56,57^. Future research will explore pleiotropic genetic effects to better understand the link between obesity and its comorbidities.

A recent study demonstrated that PRS can identify individuals at high risk of obesity. By analyzing over 2 million variants in 300,000 people, the PRS explained 8.4% of BMI variation^28^, with those in the top decile having a BMI 2.9 kg/m² higher on average, and a 4.2-fold increased risk of SevO compared to the lowest nine deciles. Our PRS analysis, using fewer variants and a smaller sample, we found a different BMI distribution at the tail ends of the distribution of genetic susceptibility, highlighting the potential for heterogeneity in the relationship between PRS and BMI distribution. We found SevO in 4.9% of individuals in the top 90th percentile vs. 0.55% in the bottom 10th percentile, indicating a 10-fold increased risk. However, our study and previous studies showed low predictive value, with AUC_ROC_ values ranging from 0.16 to 0.33 and sensitivity from 0.15 to 0.39^28,36^. This poor prediction may stem from the exclusion of lifestyle factors and selective participation bias in the UKBB. Our analysis suggests that incorporating lifestyle factors, like diet and physical activity, is crucial for improving personalized risk prediction.

There were several limitations to our study. Sample sizes were small for certain populations (e.g., East Asians) and SevO classes (Obesity Class IV). There was also a sex imbalance between the predominantly female discovery cohort and predominantly male replication cohort, limiting validation of sex-specific associations. Additionally, the 95% percentile BMI trait showed heterogeneity, lower predictability, and less heritability, which may have reduced GWAS power. Despite this, we conducted a complementary GWAS using WHO SevO classification to identify additional variants. Lastly, we recognize that stratification based on self-reported background may not fully align with genetic similarity, but it was necessary due to limitations in existing cohorts.

Despite these limitations, our study identified a novel locus influencing SevO and expands the number of identified SevO signals. We also confirmed a strong overlap between the genetic architecture of SevO and BMI. The integration of two replicated variants (located in or near *BHLHE40-AS1,* and *PLCL2,*) with transcriptomic data suggests a candidate gene in each region with the top GWAS SNP likely influencing risk of SevO through differential expression. Follow-up functional studies for these regions will be prioritized in our future work along with causal modelling to infer genetic liability. Our systematic analysis combining PRS, PheWAS and lifestyle factors further reinforced the limited etiologic heterogeneity between obesity traits, the common downstream sequelae associated with SevO and BMI, and the importance of lifestyle factors in understanding genetic risk for SevO.

## MATERIALS AND METHODS

### Study Design

Study-specific design, sample quality control and descriptive statistics are provided in **Supplementary Tables 1-3**. We conducted a two-stage study of SevO traits: 95^th^ percentile [cases defined as the upper 5^th^ percentile and controls as the 5^th^ −50^th^ percentile], Obesity Class III [BMI ≥ 40 kg/m^2^] vs. controls [BMI ≥ 25 kg/m^2^] and Obesity Class IV [BMI ≥ 50 kg/m^2^] vs. controls [BMI ≥ 25 kg/m^2^]. Stage 1 discovery analyses consisted of up to 159,359 individuals (≥18 years to < 70 years) across 11 diverse population-based studies which includes individuals of European (*n=*108,844), African (*n=*23,237), Hispanic (*n=*20,081), East Asian (*n=*4,138), American Indian (*n=*276), Hawaiian (*n=*2,232) and mixed ancestry (*n=*508) backgrounds (**Supplementary Table 2**). Of note, the numbers in the ancestry -combined samples do not match our ancestry-stratified samples because of the inclusion of datasets that did not initially meet our ancestry-stratified criteria (N in cases >20). Discovery meta-analyses were performed in each ancestry group separately and in an all-ancestry combined group for both sex-specific and sex-combined analyses. Given the preponderance of rare variations in our initial discovery, we assessed replication of our novel independent variants that reached GWAS significance threshold (*P =* 5×10^−8^) in a stage 2 follow-up samples of 480,897 individuals (≥18 years to < 70 years) across 6 diverse population-based studies which includes individuals of European (*n*=390,569), African/African American (*n=*62,539), Hispanic (*n=*21,781), South Asian (*n=*4,759) and East Asian (*n=*1,557) ancestries (**Supplementary Table 2**). All studies were approved by local research ethics committees across each institution, and all participants gave informed consent. All methods were performed in accordance with the relevant guidelines and regulations.

### Phenotype Definitions

We used 3 different classes of SevO traits defined by BMI (weight (in kg)/height (in m)^2^). Using ancestry specific quantile regression modelling nested within our cohorts, the 95^th^ percentile SevO class was defined as the upper 5th percentile (cases) and 5^th^-50^th^ percentile (controls) of BMI distribution stratified by sex after controlling for age, age^2^ and ancestry (**Supplementary Figures 1-4**). For clinical obesity classes, cases were defined by BMI ≥ 40kg/m^2^ for Obesity Class III and BMI ≥ 50kg/m^2^ for Obesity Class IV. Controls were subjects with BMI ≥ 18.5kg/m^2^ and < 25kg/m^2^. A minimum of 20 cases and 20 controls for each study-specific stratum was required for combined analyses, whereas a minimum of 10 cases and 10 controls for each study-specific stratum was required for sex-specific analyses.

### Association analyses

Ancestry and sex specific GWAS, adjusted for age, principal components of ancestry (PCs) 1-10 and study specific covariates, were conducted for the three different classes of SevO traits. Poorly imputed variants (IMPUTE info < 0.4 and/or *R*^2^ < 3)^58^, and those with an effective sample size less than 20 in each stratum (or 10 in each sex-specific stratum) were excluded from association analyses. A centralized quality control procedure implemented in EasyQC^58^ was applied to association summary statistics to identify study specific problems. Ancestry and sex combined meta-analysis for each SevO trait was performed in METAL^59^ using the fixed-effect inverse variance method based on β estimates and standard errors from each study. Variants with a minor allele count [MAC] <50, with a combined sample size of <100, and an inflated heterogeneity test score (Het *I*^2^) > 80 (for one study) were excluded from downstream analyses. Similar GWAS, meta-analysis and quality control methods were employed for the replication analyses.

### Gene mapping, functional annotation, and validation of novel variants

Post-GWAS analysis of gene mapping, functional annotation, and tissue expression analysis of prioritized loci in our discovery GWAS was conducted using Functional Mapping and Annotation (FUMA)^60^ SNP2GENE function. We identified each locus with an index SNP that met genome-wide significant *P* value threshold of *P* 5×10^−8^ and then included all SNPs surrounding the index SNP ±250kb on each side. We functionally annotated each locus by considering the index SNP and any SNP in the 500kb region that displayed linkage disequilibrium (LD) *r*^2^<0.6 with the index SNP using the phase all ancestry 1000G project LD reference panel. If in some loci, two or more SNPs achieved genome-wide significant evidence for association but were independent of one another (*r*^2^ <0.1), we functionally annotate each independent SNP effect. If no genes were present within ±250 kb of the lead variant, the closest gene was selected. We also used PhenoScanner v2^61^, GWAS Catalog^62^, and the Integrative Epidemiology Unit (IEU) Open GWAS Project, as well as conducting a comprehensive literature review to evaluate our independent associated variants across all three SevO classes. A variant is considered novel if not previously associated with obesity related traits. Variants that meet these criteria were subsequently assessed in our replication dataset and were considered significant if they were directionally consistent and met our Bonferroni adjusted significance threshold (0.05/*n* independent SNPs) in any SevO class. Independent unreported variants were annotated for predicted pathogenicity by Combined Annotation Dependent Depletion (CADD) scores using CADDv1.3^63^ with a Phred-scale CADD score >10 (top 10%) being deleterious. We utilized online resources such as GeneCards, GWAS Catalog and PubMed to derive potential biological links of genes in proximity of novel signals (±250kb of the index SNP, or the closest gene if no gene was present within ±250 kb) (**Supplementary Table 12**).

### Expression Quantitative Trait Loci (eQTL) and colocalization

We used eQTL summary data from GTEx v8^26^ and eQTLGen Phase 1 release^27^, to test for *cis* associations between our novel findings with transcripts within ±1 Mb across obesity related tissues including liver, skeletal muscle, whole blood, brain, and adipose tissues surrounding the transcription start (TSS). Detailed method descriptions can be found in the main GTEx v8^26^ and/or eQTLGen^27^ publications. Additionally, we examined *cis* associations between our SevO novel variants with transcripts derived from RNA sequencing (RNAseq) within ±1 Mb in whole blood among 645 Mexican Americans from the County Hispanic Cohort (CCHC)^64^. Significant eQTLs were determined based on Bonferroni adjusted significance threshold (0.05/*n* of transcripts tested for the locus). Colocalization of any validated novel variants in the CCHC cohort was performed using the *coloc* software^65^ with a posterior probability >75% supporting colocalization from a single causal variant. Detailed method descriptions on CCHC study population, genotyping and eQTL mapping are provided in Chen *et al*. 2022^43^

### Comparison of Replicated SNPs With Previous Publication

We tested the direction of association of in our discovery analysis for Obesity Class III with 813 known BMI-associated variants (based on HapMap imputed data) that met a significance threshold of *P<*5 × 10^−05^, from approximately 700,000 European participants identified in Yengo *et al* ^12^. We also performed lookups of our unreported independent validated variants (or their LD proxies: R^2^ >0.8) with SevO traits within this cohort to assess consistency of associations in both the meta-analysis of GIANT and UKBB^12^ and the UKBB trans-ancestry meta-analysis of BMI.

### SNP based heritability’s and genetic correlation between SevO and BMI

Using GWAS summary statistics, we calculated single nucleotide polymorphisms (SNP)-based heritability for SevO traits and the genetic correlation between each trait with BMI using linkage disequilibrium score regression (LDSC)^70,71^. Heritability, ranging from 0 to 1, measures the proportion of variation in a phenotype accounted for by genetic factors. SNP-based genetic correlations, ranging from −1 to +1, measure the extent to which two phenotypes share common genetic variation. Analyses were conducted in the ancestry -combined samples and in European -combined samples only.

### Polygenic Risk Score (PRS)

Polygenic risk scores (PRS) were generated using GWAS summary statistics from three comparative sets: a) the significant lead independent SNPs from each SevO trait all ancestry GWAS, b) the HapMap 3 SNPs in the all ancestry GWAS of SevO traits from the current study, and c) the HapMap 3 SNPs in the BMI GWAS from the Locke *et al* (2015) paper in the GIANT consortium^14^. Both the UKBB and CCHC test cohorts were not included in the generation of the PRS. We utilized the PRS-continuous shrinkage (CS) method which applies a Bayesian regression framework and places conceptually CS priors on variant effect estimates^72^. Pairwise LD matrices within pre-defined LD blocks^73^ (using European LD blocks for LD calculations were calculated using PLINK and converted to HDF5 format^74^). PRS for SevO or BMI was calculated for each ancestral group across all SevO classes. Multivariable logistic regression was used to test the association of PRS SevO or BMI with SevO classes adjusted for age, sex, 10 PCs and ancestry in the UKBB. A partial Nagelkerke *R*^2^ was used to estimate the proportion of variance for SevO classes explained by the PRS.

### PheWAS using the UKBB

Using the weighted Obesity Class III PRS-BMI we performed a PheWAS of 19 clinical classes of traits in the UKBB for each ancestry (African, European, South Asian, and East Asian). Detailed description of the classes of clinical traits are described in **Supplementary Table 16.** Regression modelling with Obesity Class III PRS-BMI as the independent variable, phecodes as the dependent variables, and age, sex, the first 10 PCs as covariates, were used to identify phenotypic associations. An FDR of 0.05 was applied to account for multiple testing.

### Measures of Diagnostic Accuracy

Sensitivity, Specificity, Positive Predictive Values (PPV), Negative Predictive Values (NPV) and area under the curve (AUC) was applied in evaluating the effectiveness of SevO PRS only, covariates only and combined SevO PRS and covariates with Obesity Class III across European, African, and South Asian participants from the UKBB and Hispanic participants from the CCHC cohort.

### PRS deciles and assessment of lifestyle factors

We stratified UKBB participants by their Class III SevO PRS-BMI according to deciles (1-10) with high PRS-BMI comprising >90^th^ percentile (decile 10) and low PRS-BMI the ≤10^th^ percentile (decile 1). Normal weight (≤25 kg/m^2^), overweight (>25 kg/m^2^ ≤30 kg/m^2^) obesity (>30 kg/m^2^ ≤40 kg/m^2^) and SevO Class 3 (>40 kg/m^2^) was determined within each decile for ancestry -stratified and -combined groups. We explored the predictive power of our PRS-BMI on severe obesity (≥ 40 kg/m^2^) between our > 90^th^ percentile (*n=* 42,844) and ≤10^th^ percentile (*n=* 42,856) in the ancestry -combined samples within UKBB. We then assessed whether patterns of lifestyle factor associations, including physical activity, dietary patterns, alcohol/smoke servings, sleep behaviors and self-reported “body size” at age 10, were differentially associated in individuals across the PRS-BMI deciles. Those with a PRS in the ≥ 90^th^ percentile and a BMI < 25 kg/m^2^ were labeled the “Resilient group”. Lifestyle behavior regression modelling was compared between our “Resilient group” and those with 1) PRS and SevO; 2) >90^th^ percentile for PRS and SevO Class III; 3) < 10^th^ percentile for PRS and SevO and 4) <10^th^ percentile and BMI <25 kg/m^2^.

## Supporting information

Author Contributions

Funding

Studies

## Data availability

The datasets generated and/or analyzed during the current study will be available in the GWAS Catalog repository.

