## Supplementary Figures for "Genome-wide association study provides novel insight into the genetic architecture of severe obesity"

**Supplementary Figure 1.** Women, standard cut-offs versus by quantile regression. Jagged lines for %tile cut-offs are because all races are shown together.

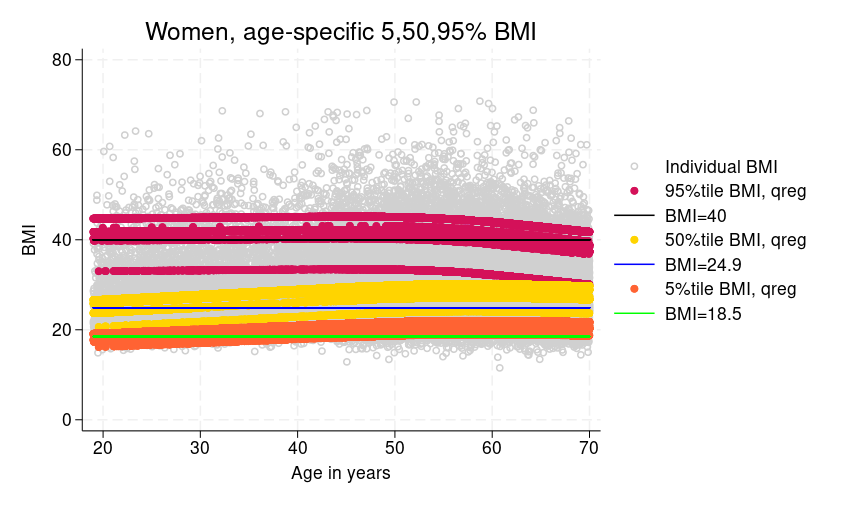

**Supplementary Figure 2.** Men, standard cut-offs versus by quantile regression. Jagged lines for %tile cut-offs are because all races are shown together.

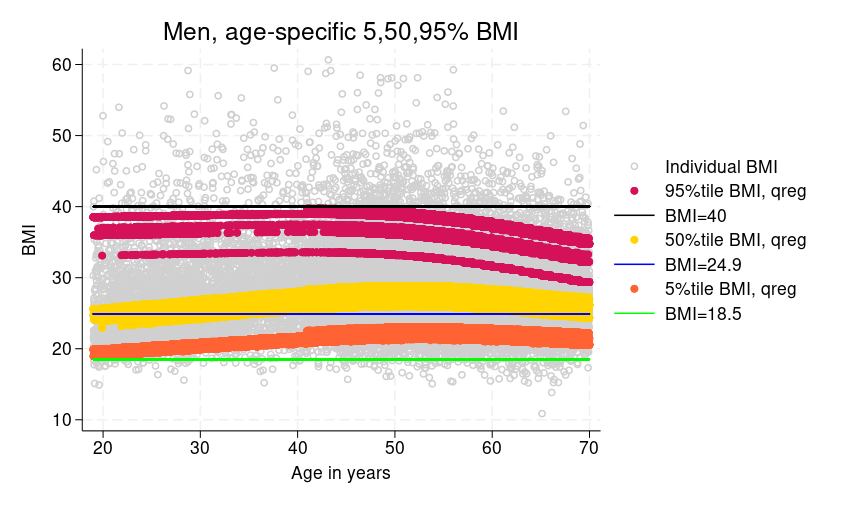

**Supplementary Figure 3.** Women, quantile regression by race/ethnicity.

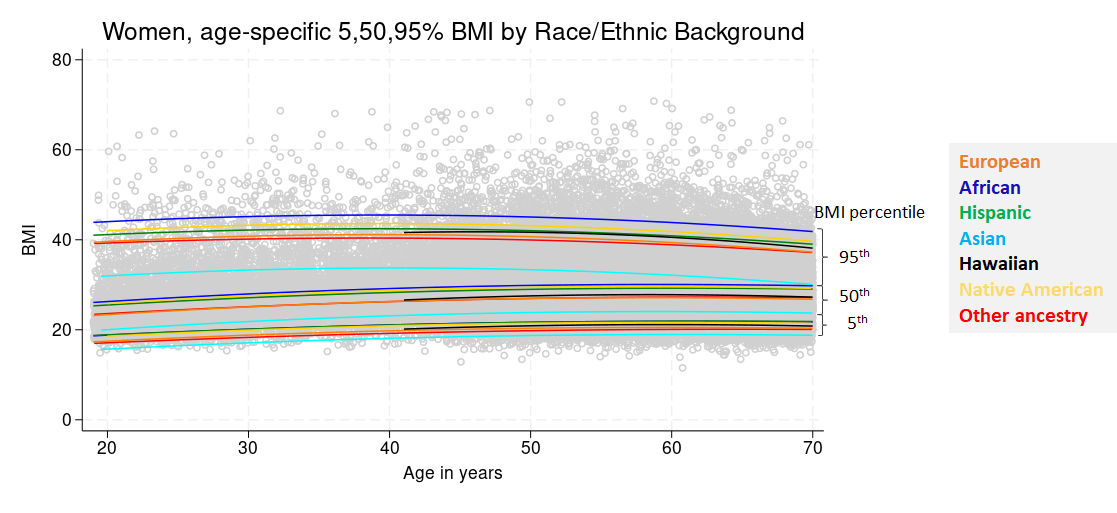

**Supplementary Figure 4.** Men, quantile regression by race/ethnicity.

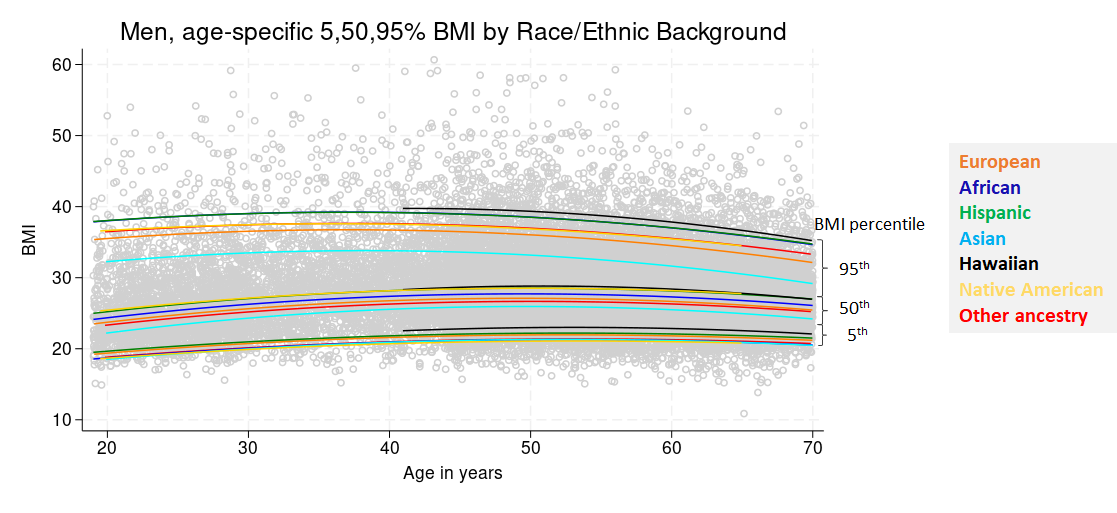

**Supplementary Figure 5.** QQ (upper) and Manhattan (lower) plots for All-ancestry sex combined 95th percentile severely obese vs 5th to 50th percentile normal weight controls. Black dots in QQ plot denote expected vs observed -log 10 p-values for association without controlling for known obesity loci, while dark orange dots denote expected vs observed after controlling for known obesity loci. Blue dots in Manhattan plot denote known signals and red dots suggest possible novel signals.

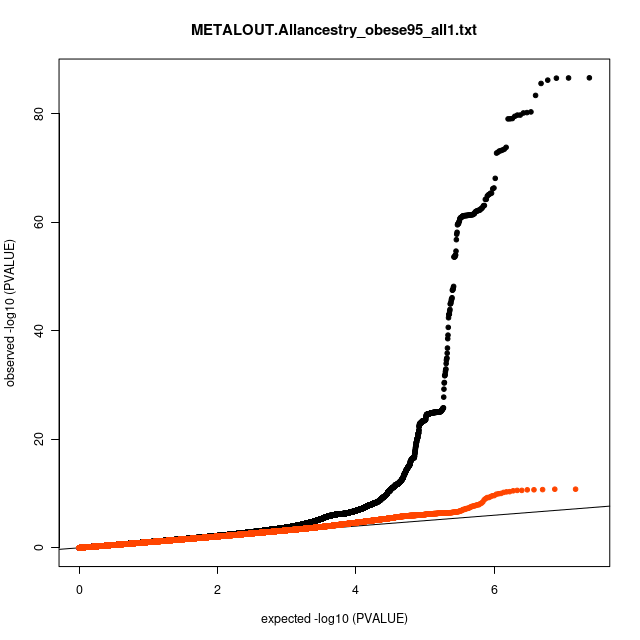

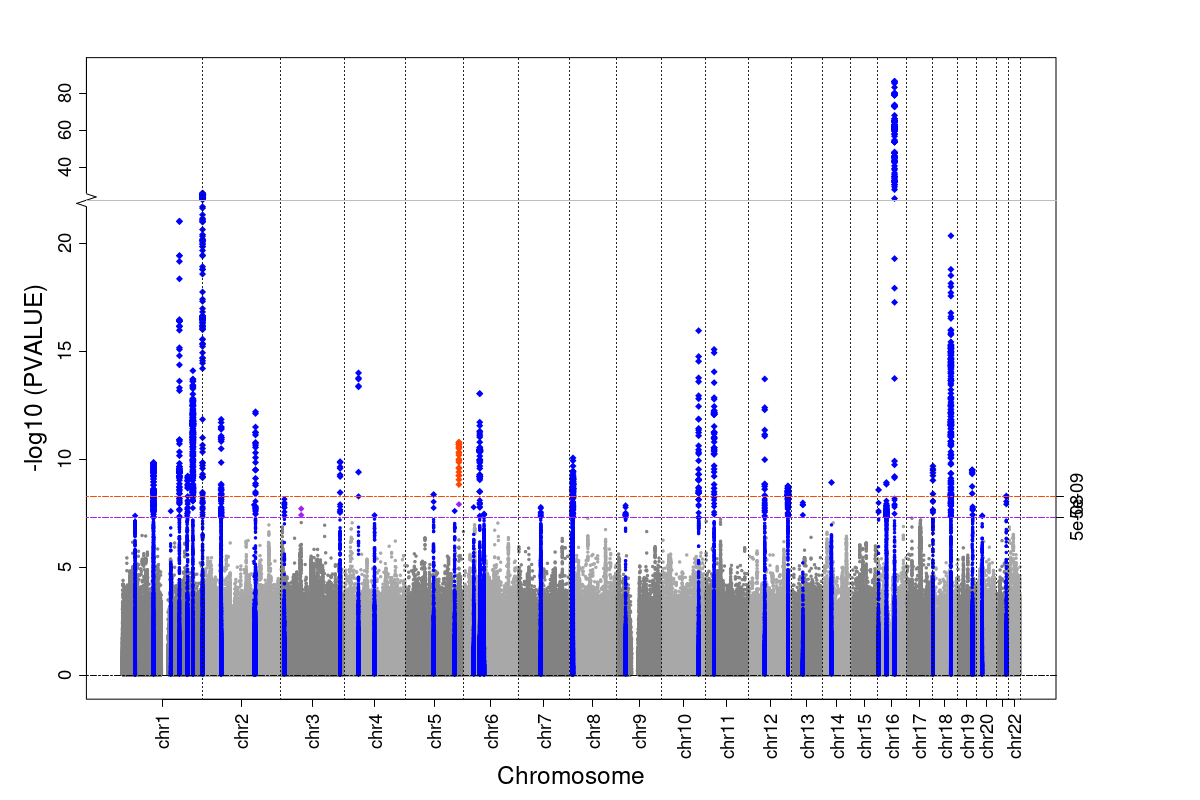

**Supplementary Figure 6.** QQ (upper) and Manhattan (lower) plots for All-ancestry female 95th percentile severely obese vs 5th to 50th percentile normal weight controls. Black dots in QQ plot denote expected vs observed -log 10 p-values for association without controlling for known obesity loci, while dark orange dots denote expected vs observed after controlling for known obesity loci. Blue dots in Manhattan plot denote known signals and red dots suggest possible novel signals.

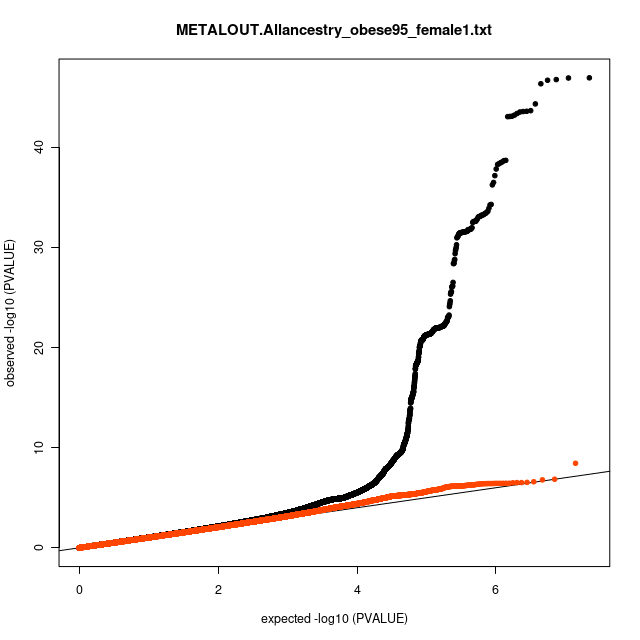

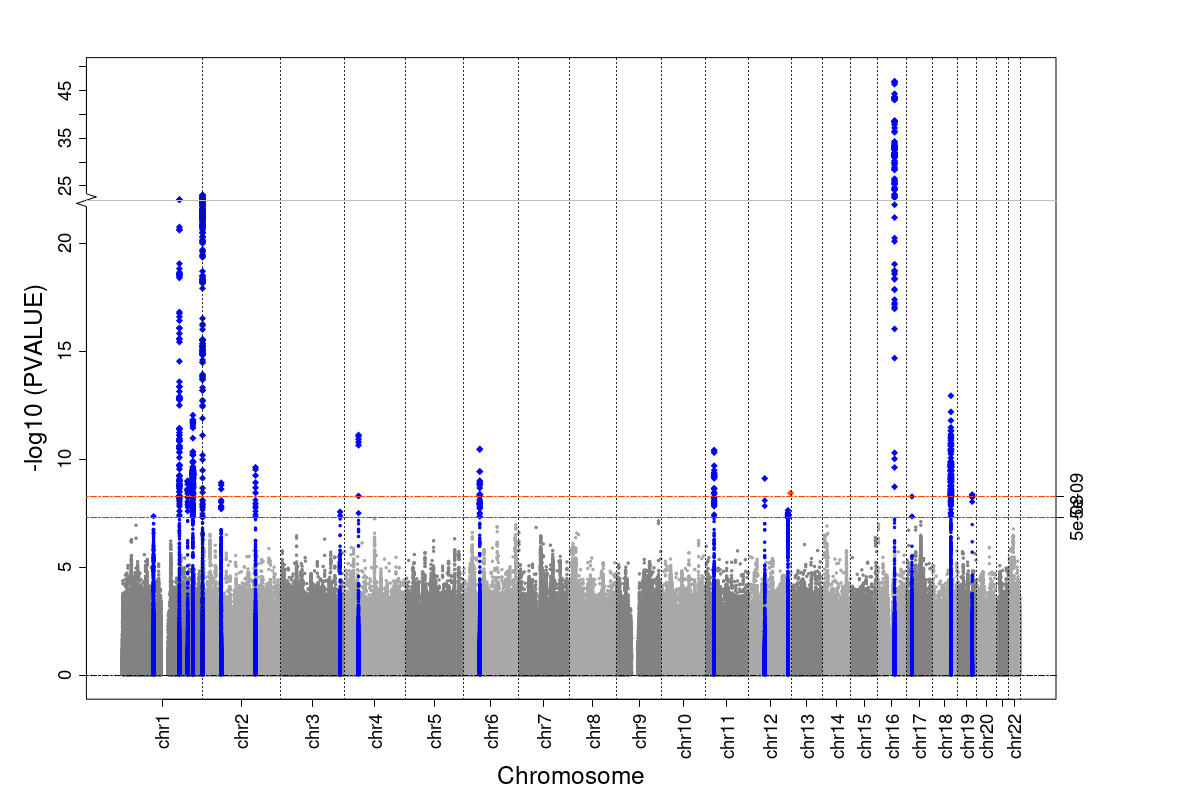

**Supplementary Figure 7.** QQ (upper) and Manhattan (lower) plots for All-ancestry male 95th percentile severely obese vs 5th to 50th percentile normal weight controls. Black dots in QQ plot denote expected vs observed -log 10 p-values for association without controlling for known obesity loci, while dark orange dots denote expected vs observed after controlling for known obesity loci. Blue dots in Manhattan plot denote known signals and red dots suggest possible novel signals.

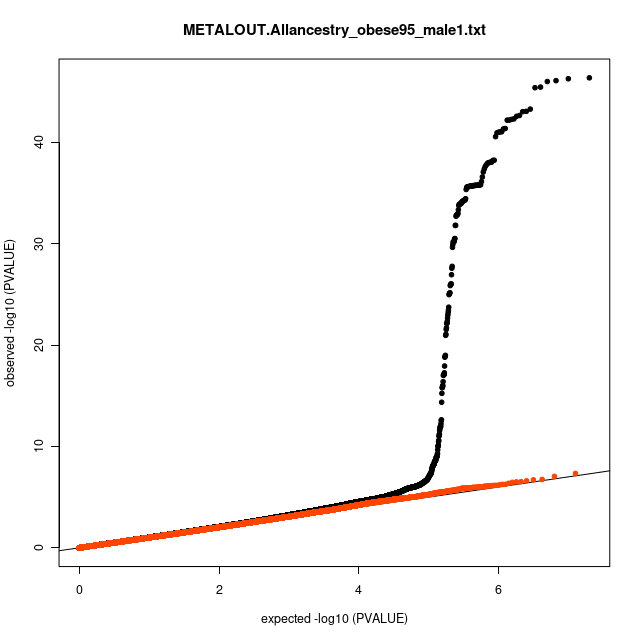

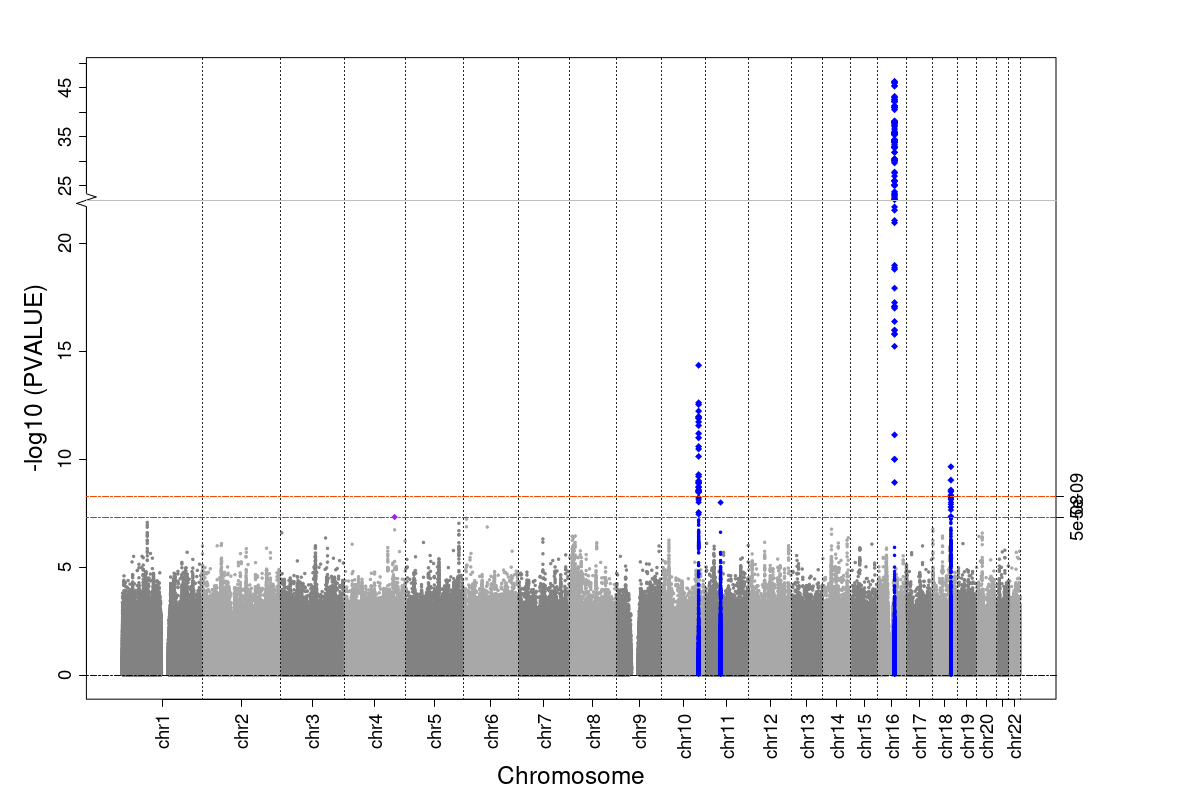

**Supplementary Figure 8.** QQ (upper) and Manhattan (lower) plots for All-ancestry sex combined class III (BMI≥40 kg/m^2^) vs normal weight (18 kg/m^2^≤BMI<25 kg/m^2^) controls. Black dots in QQ plot denote expected vs observed -log 10 p-values for association without controlling for known obesity loci, while dark orange dots denote expected vs observed after controlling for known obesity loci. Blue dots in Manhattan plot denote known signals and red dots suggest possible novel signals.

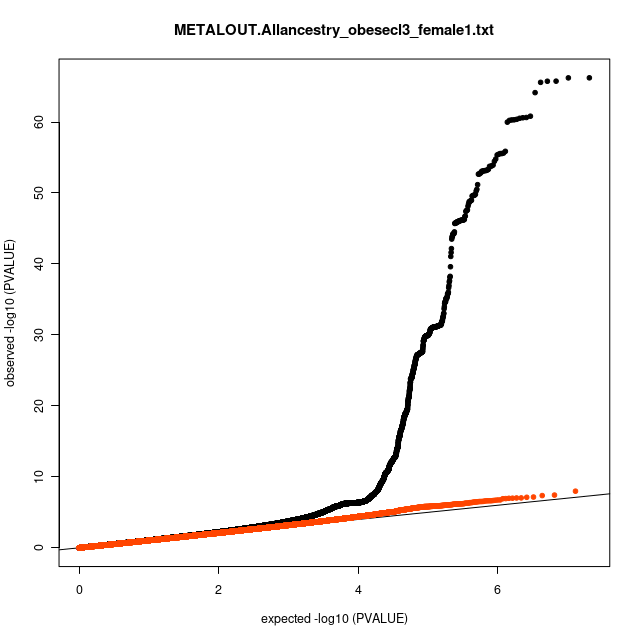

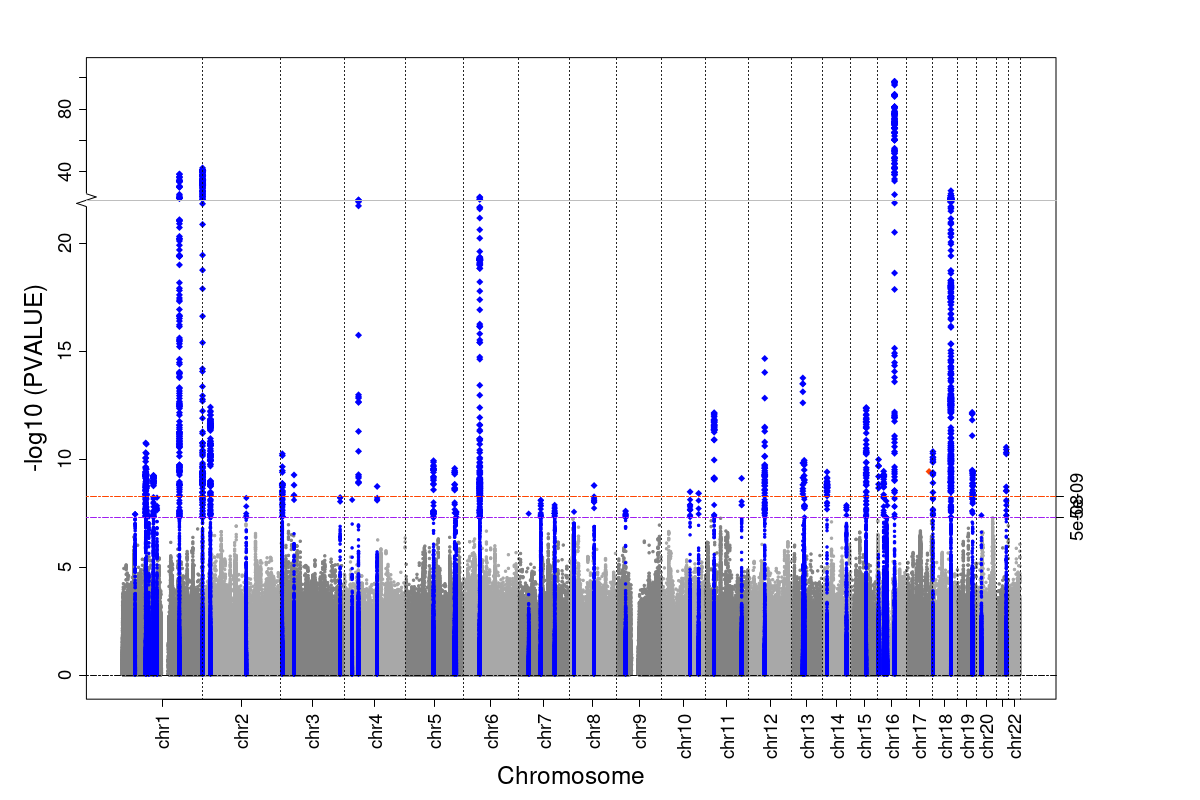

**Supplementary Figure 9.** QQ (upper) and Manhattan (lower) plots for All-ancestry female class III (BMI≥40 kg/m^2^) vs normal weight (18 kg/m^2^≤BMI<25 kg/m^2^) controls. Black dots in QQ plot denote expected vs observed -log 10 p-values for association without controlling for known obesity loci, while dark orange dots denote expected vs observed after controlling for known obesity loci. Blue dots in Manhattan plot denote known signals and red dots suggest possible novel signals.

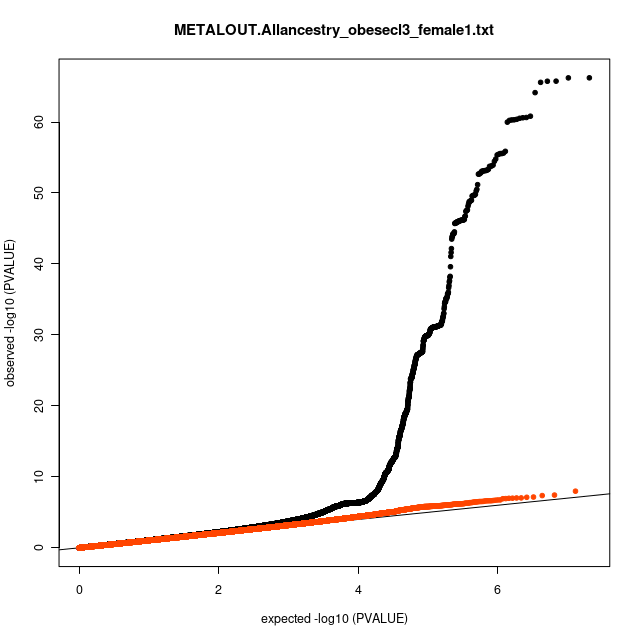

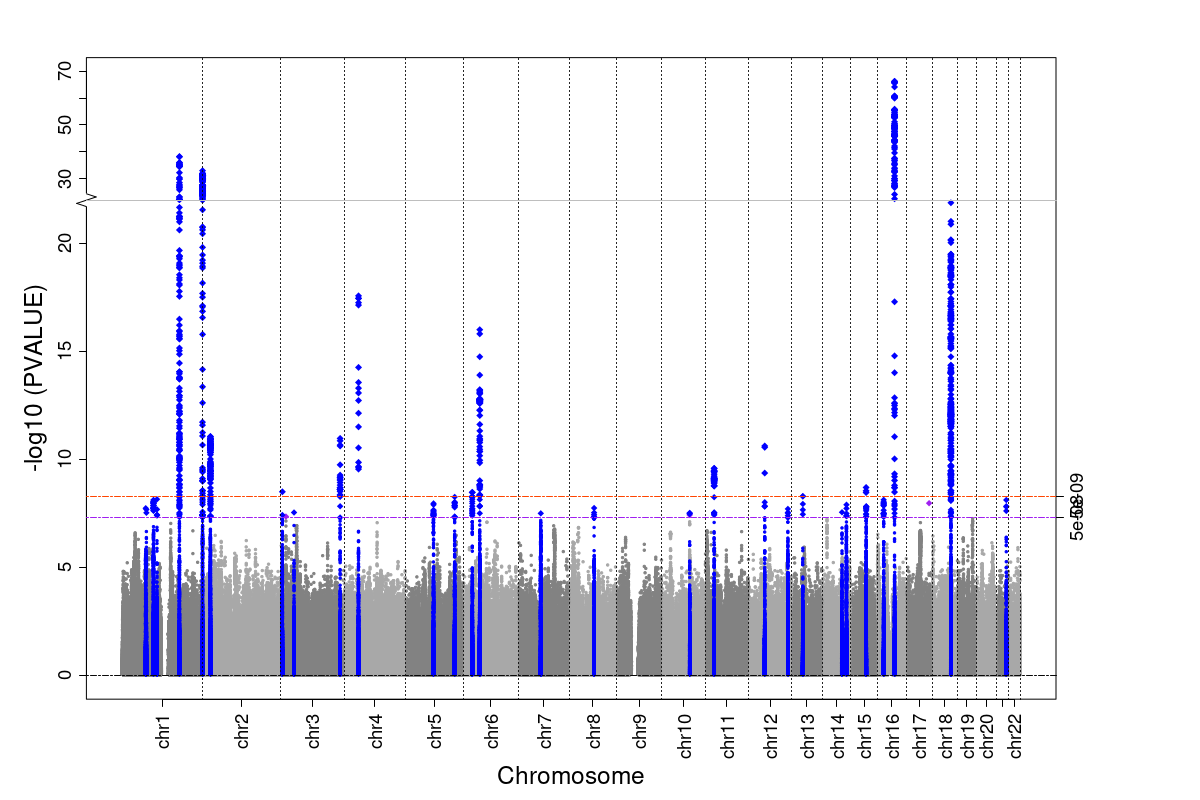

**Supplementary Figure 10.** QQ (upper) and Manhattan (lower) plots for All-ancestry male class III (BMI≥40 kg/m^2^) vs normal weight (18 kg/m^2^≤BMI<25 kg/m^2^) controls. Black dots in QQ plot denote expected vs observed -log 10 p-values for association without controlling for known obesity loci, while dark orange dots denote expected vs observed after controlling for known obesity loci. Blue dots in Manhattan plot denote known signals and red dots suggest possible novel signals.

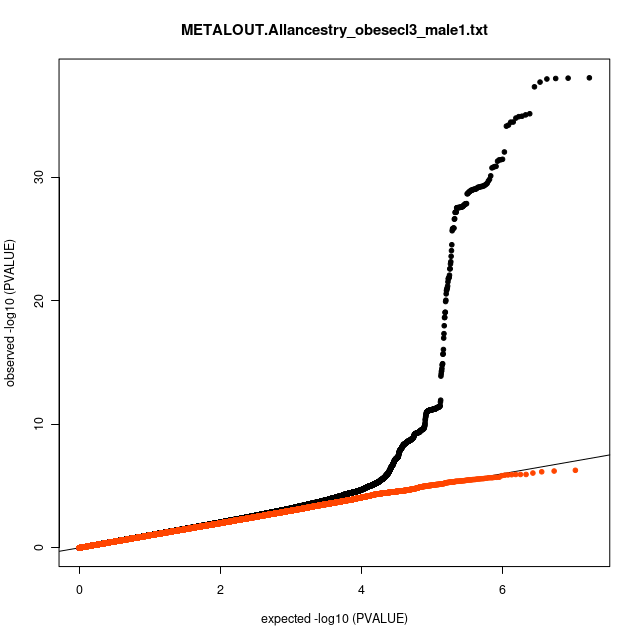

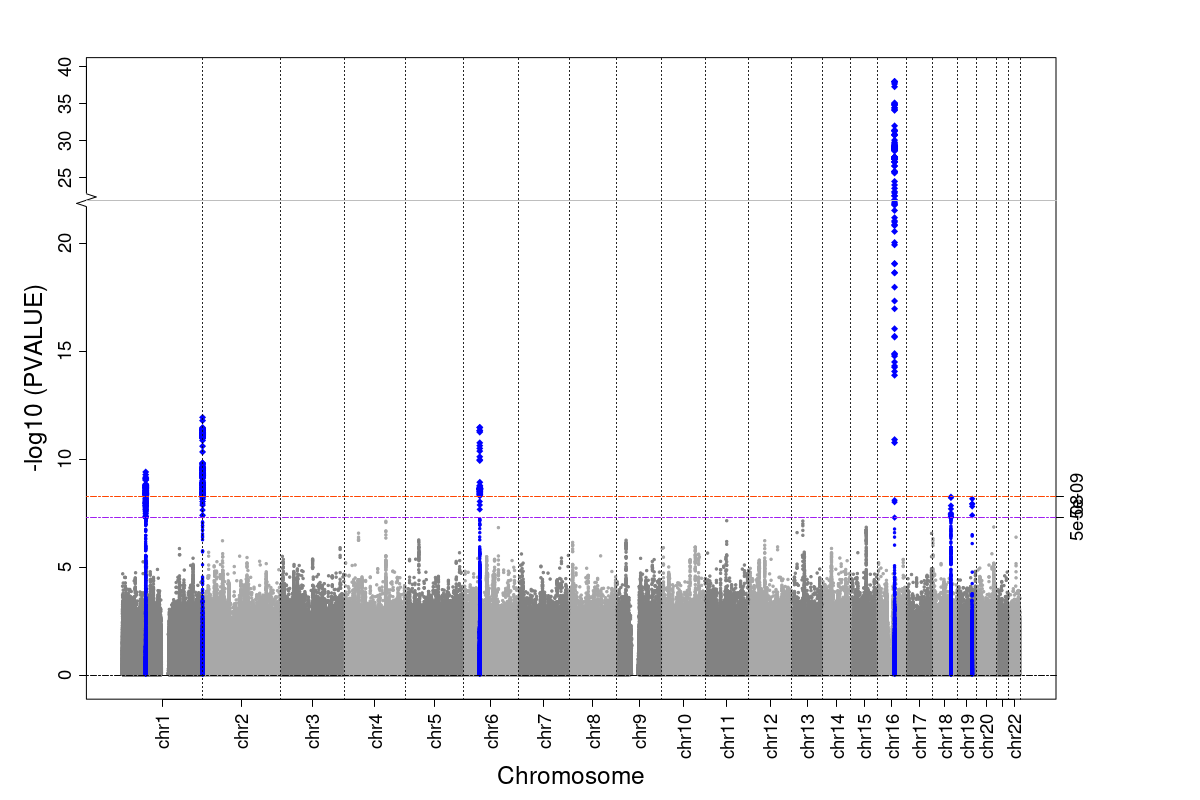

**Supplementary Figure 11.** QQ (upper) and Manhattan (lower) plots for All-ancestry sex combined class IV (BMI≥50 kg/m^2^) vs normal weight (18 kg/m^2^≤BMI<25 kg/m^2^) controls. Black dots in QQ plot denote expected vs observed -log 10 p-values for association without controlling for known obesity loci, while dark orange dots denote expected vs observed after controlling for known obesity loci. Blue dots in Manhattan plot denote known signals and red dots suggest possible novel signals.

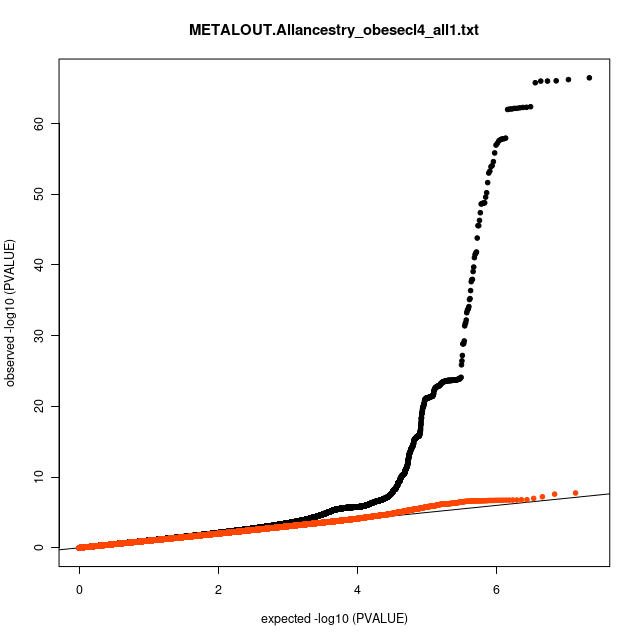

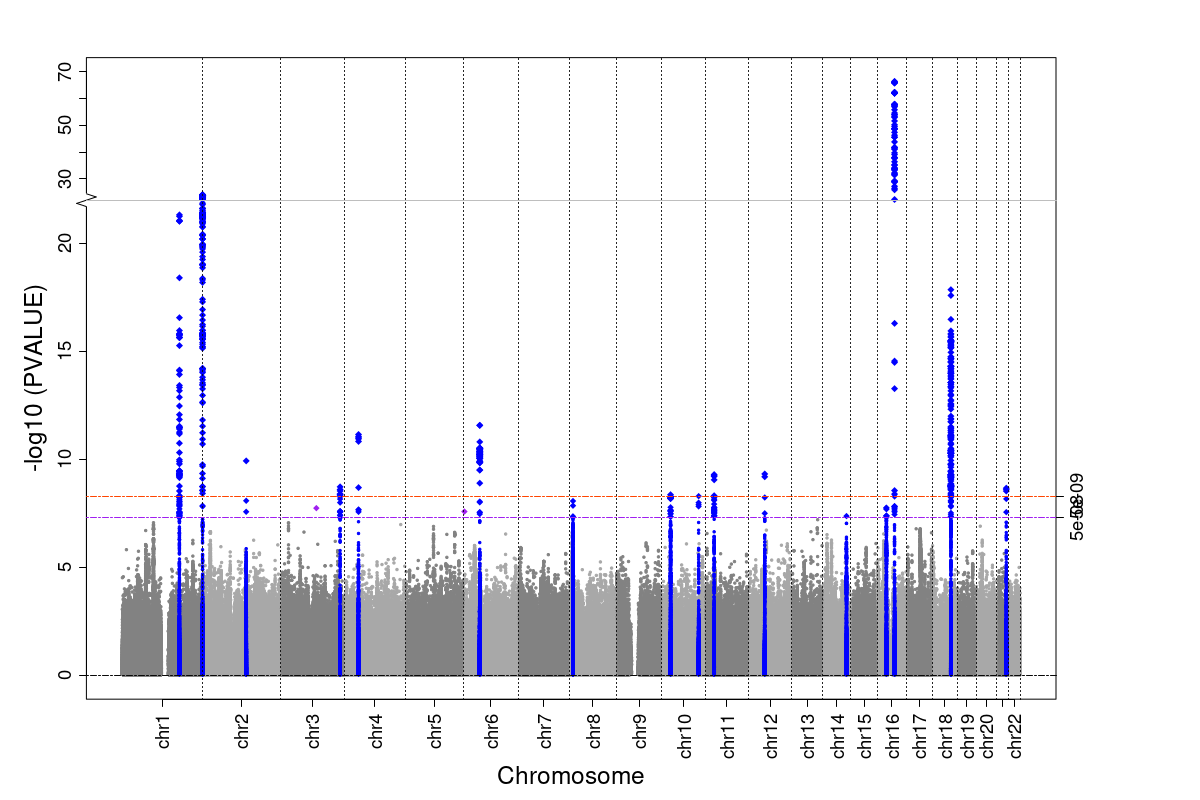

**Supplementary Figure 12.** QQ (upper) and Manhattan (upper) plots for All-ancestry female class 4 (BMI≥50 kg/m^2^) vs normal weight (18 kg/m^2^≤BMI<25 kg/m^2^) controls. Black dots in QQ plot denote expected vs observed -log 10 p-values for association without controlling for known obesity loci, while dark orange dots denote expected vs observed after controlling for known obesity loci. Blue dots in Manhattan plot denote known signals and red dots suggest possible novel signals.

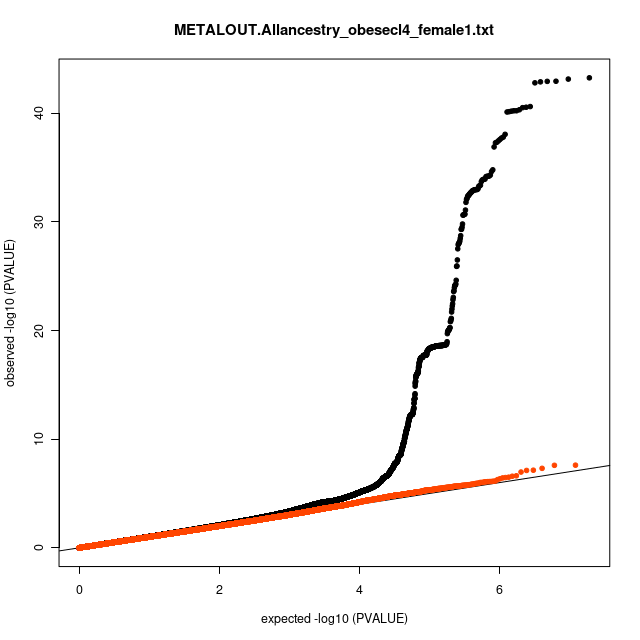

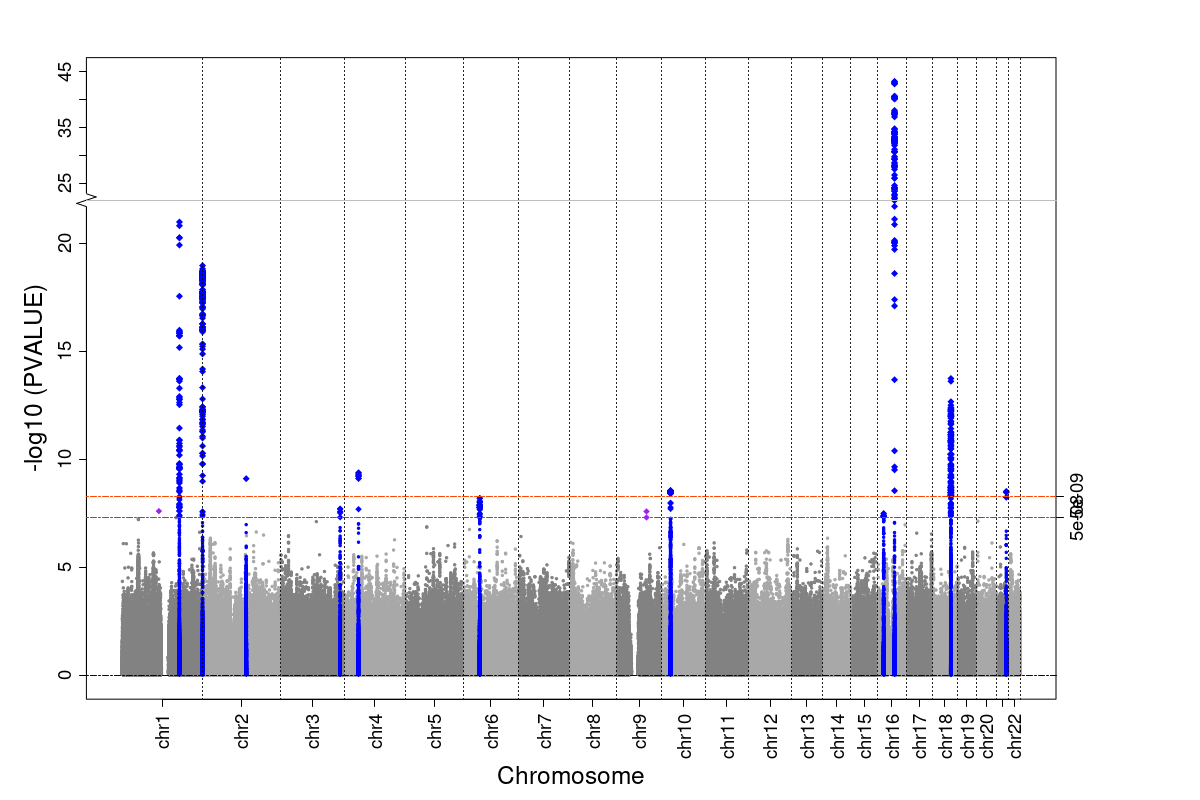

**Supplementary Figure 13.** QQ (upper) and Manhattan (lower) plots for All-ancestry female class 4 (BMI≥50 kg/m^2^) vs normal weight (18 kg/m^2^≤BMI<25 kg/m^2^) controls. Black dots in QQ plot denote expected vs observed -log 10 p-values for association without controlling for known obesity loci, while dark orange dots denote expected vs observed after controlling for known obesity loci. Blue dots in Manhattan plot denote known signals and red dots suggest possible novel signals.

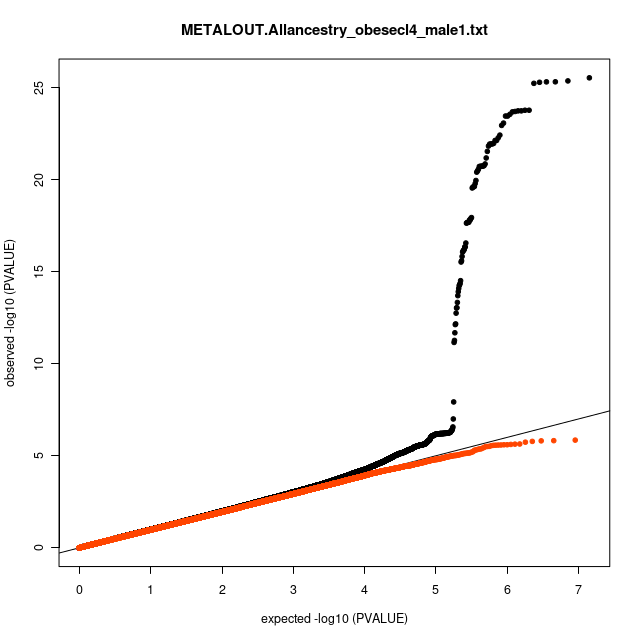

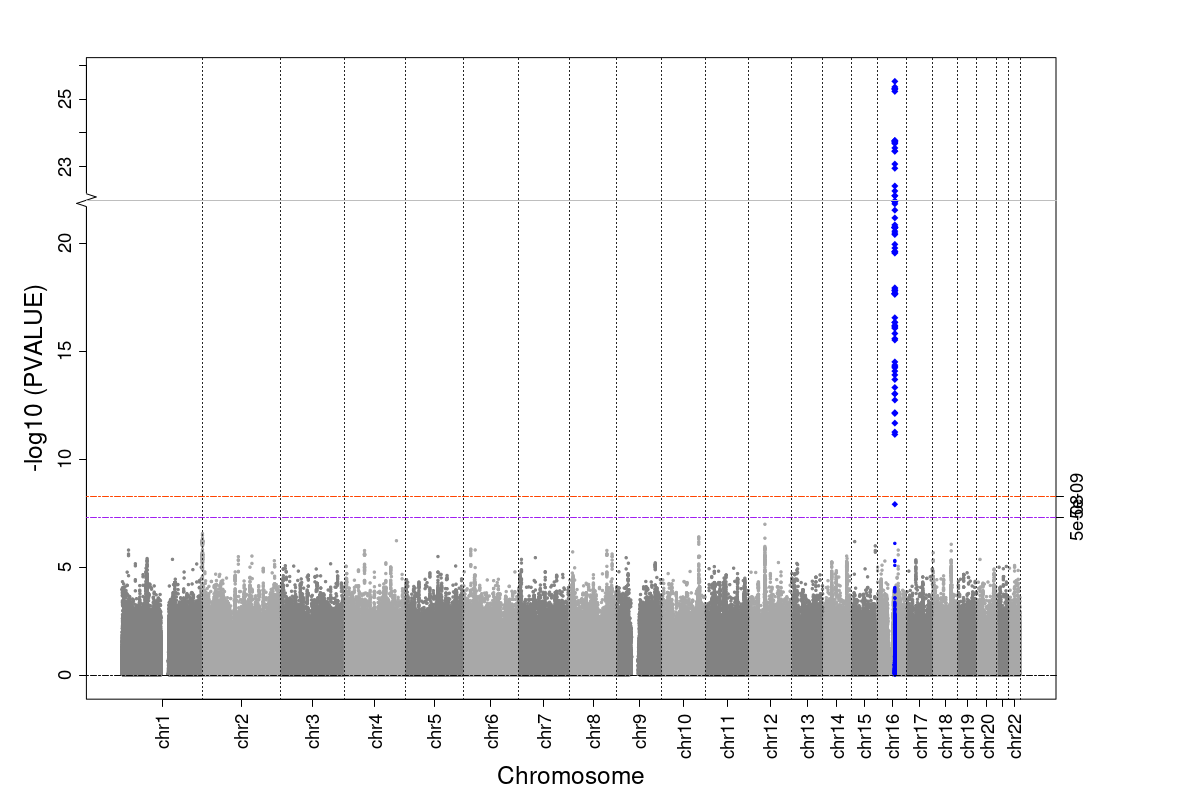

**Supplementary Figure 14.** Phenome-wide association of severe Obesity Class III polygenic risk score with clinical traits in self-identified East Asian, African, and South Asian ancestry groups.

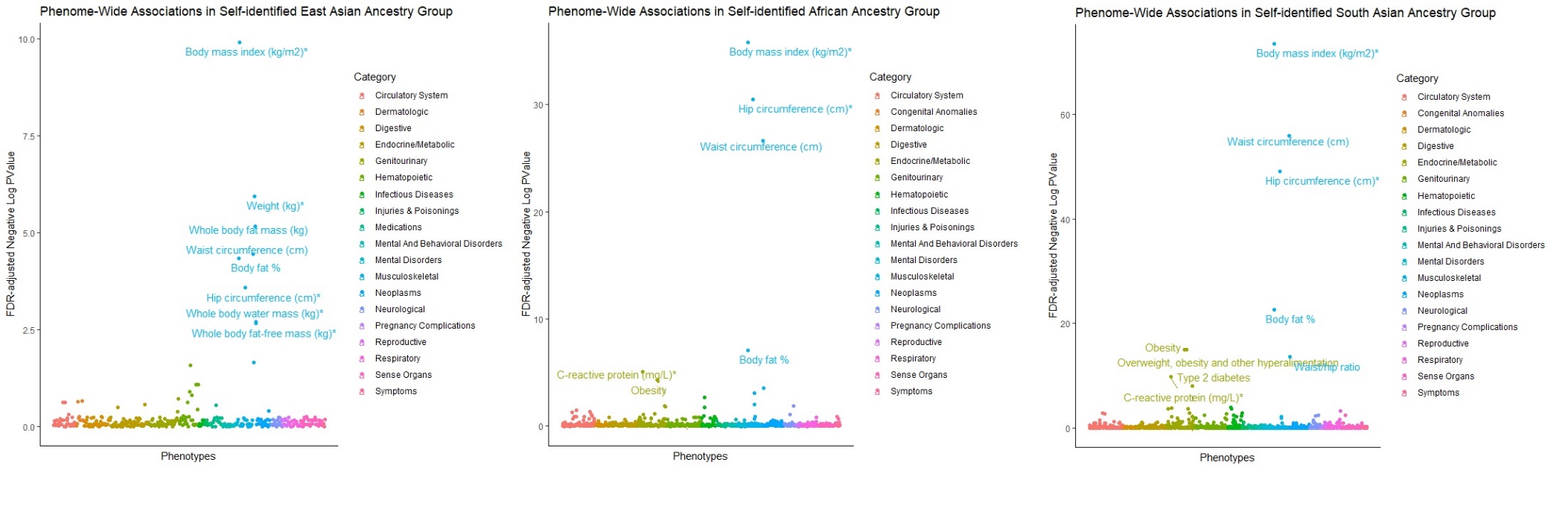

**Supplementary Figure 15.** Comparison of direction and effect sizes of replicated severe Obesity Class III associated SNPs (at GWAS significance level) with summary statistics of the same SNPs from GIANT BMI GWAS meta-analyses. Severe obesity beta represents log odds and may not be likewise comparison to betas reported in GIANT. However, results suggest significant consistency in both direction and relative effect sizes.

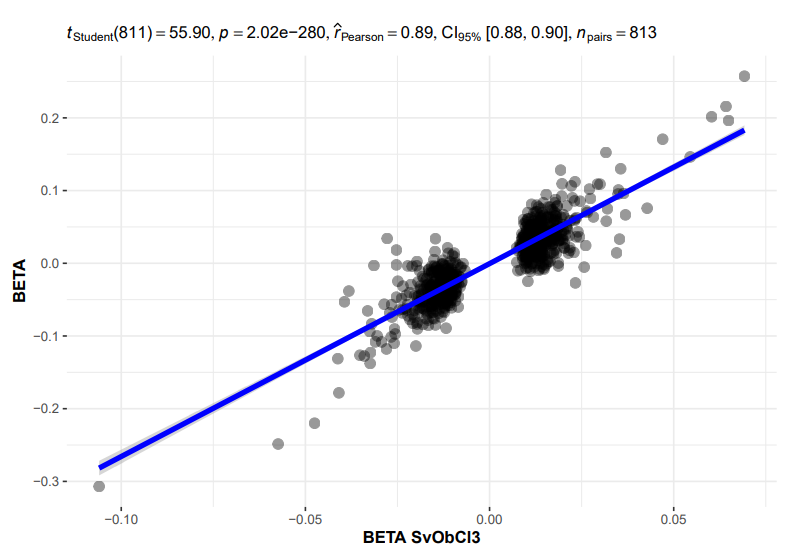

**Supplementary Figure 16.** BMI distributions by self-identified ancestry groups comparing those in PRS>90^th^ percentile versus individuals in <10^th^ percentile category. African and East Asian ancestry groups in the lower 10^th^ percentile group show noticeably higher and lower BMI distributions compared to other ancestries respectively. East Asians show lower average BMI distributions among those in high 90^th^ percentile category, but Africans demonstrate similar BMI patterns to other ancestries. (Note: AFR (African ancestry), EAS (East Asian ancestry), EUR (European ancestry), and SAS (South Asian ancestry))

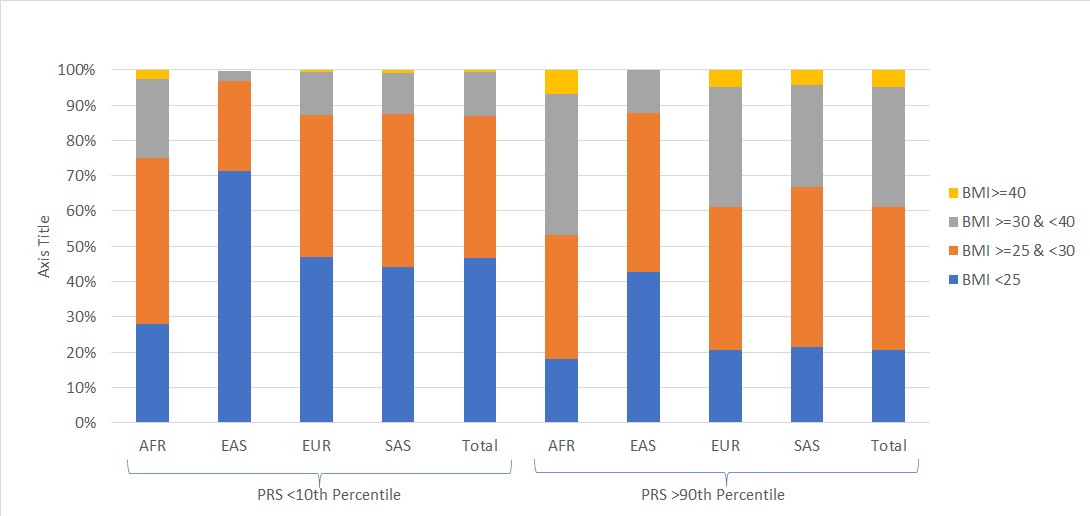

**Supplementary Figure 17.** Box plots of BMI across PRS deciles in the UKBB by ancestry background. Upward, linear association can be observed between PRS deciles and BMI in all populations. (Note: AFR (African ancestry), EAS (East Asian ancestry), EUR (European ancestry), and SAS (South Asian ancestry))

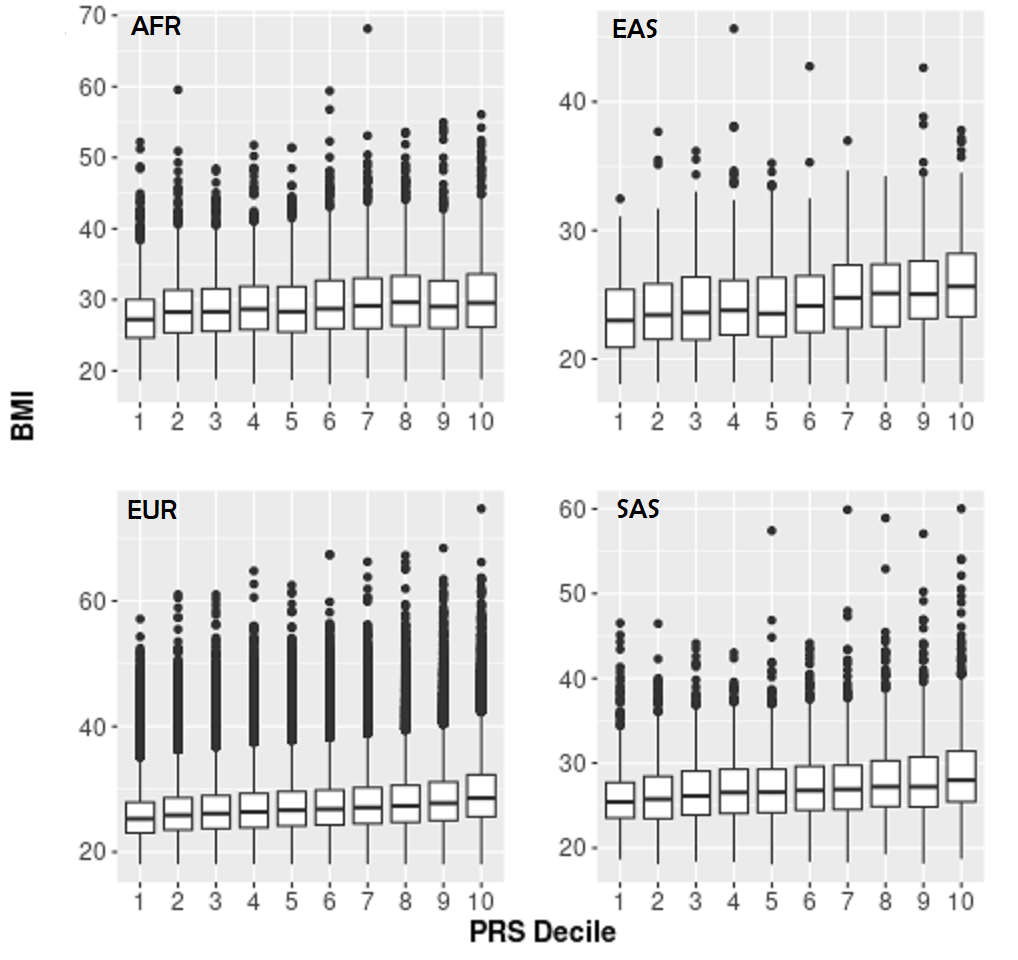
