## Supplementary material for "Genome-wide association study provides novel insight into the genetic architecture of severe obesity": Author Contributions

**Author contributions for final manuscript:**

Mariaelisa Graff and Kari E North planned the study. Mohanraj Krishnan, Mohammad Yaser Anwar, Shuwei Liu, Benjamin S Hadad and Mariaelisa Graff conducted meta and secondary analyses. Mohanraj Krishnan, Mohammad Yaser Anwar, Mariaelisa Graff, Penny Gordon-Larsen and Kari E North drafted the manuscript. All authors contributed to the discussion, critically appraised the manuscript and approved the final version for publication.

**Study specific contributions:**

**Discovery studies**

*Atherosclerosis Risk in Communities Study (ARIC)*

Heather L Highland, Kristin L Young, Marielisa Graff and Kari E North oversaw the consortium. Mohammad Yaser Anwar, Mohanraj Krishnan, Baiyu Qi and Yujie Wang performed the primary genetic analyses for the ARIC study.

*The BioMe^TM^ Biobank Program (BioMe)*

Michael H Preuss, Ryan W Walker, Eimear E Kenny and Ruth J.F Loos oversaw the consortium, managed the data and performed the primary genetic analyses for the BioMe study.

*The Coronary Artery Risk Development in Young Adults Study (CARDIA)*

Myriam Fornage led the CARIDA component of this study and prepared the genetic data for secondary analyses.

*The Multiethnic Cohort Study (MEC)*

Christopher Haiman and Ioana Cheng oversaw the consortium and performed the primary genetic analyses for the MEC study.

*Mexico Studies (MC1 & MC2)*

Esteban J Parra, Miguel Cruz, Frida Lona-Durazo, Jesus Peralta and Jamie Gomez-Zamudio oversaw the consortium, managed the data, harmonized phenotypes and performed the primary genetic analyses for the MC1 and MC2 study.

*The Hispanic Community Health Study/Study of Latinos (HCHS/SOL)*

Heather M Highland, Kristin L Young, Marielisa Graff and Kari E North oversaw the consortium. Mohammad Yaser Anwar, Mohanraj Krishnan, Baiyu Qi and Yujie Wang performed the primary genetic analyses for the HCHS/SOL study.

*The Women’s Health Initiative Study (WHI)*

Ulrike Peters and Charles Kooperberg oversaw the consortium, managed the data, harmonized phenotypes and performed the primary genetic analyses for the WHI study.

*Framingham Heart Study (FHS)*

Xiaou Zhang and Ching-Ti Liu led the data management and primary analyses of the FHS study.

*Netherlands Epidemiology of Obesity (NEO)*

Roelof A.J Smit and Renée de Mutsert oversaw the consortium, managed the data and performed the initial GWAS analyses for the NEO study.

*BIOVu*

Hung-Hsin Chen, Rashadeh Roshani and Jennifer E Below oversaw the consortium, managed the data and performed the initial GWAS analyses for the BIOVu study.

*MyCode/DiscovEHR*

Geetha Chittoor, Anne E Justice, Navya S Josyula and Christopher R Bauer oversaw the consortium, managed the data, harmonized the phenotypes and performed the primary GWAS analyses for the MyCode/DiscovEHR study.

**Replication studies**

*Million Veterans Program (MVP)*

Peter W.F Wilson and Kelly Cho oversaw the consortium and managed the data. Qin Hui, Gregorio V Linchangco, Yan V Sun, Michael J Gaziano and Luc Djousse contributed to data management and primary analysis of the MVP cohort.

*Reasons for Geographic and Racial Differences in Stroke (REGARDS)*

Mariah C Meyer and Leslie A Lange oversaw the consortium, managed the data and performed the primary analyses for the REGARDS study.

*Genetic Studies of Atherosclerosis Risk (GeneSTAR)*

Lisa R Yanek and Dhananjay Vaidya oversaw the consortium, managed the data and performed the primary analyses for the GeneSTAR study.

*Health and Retirement Study (HRS)*

Wei Zhao, Jennifer A Smith, Sharon L.R Kardia, Jessica D Faul and David R Weir oversaw the consortium, managed the data, harmonized the phenotypes and performed the primary GWAS analyses for the HRS study.

*Genetic Epidemiology Network of Arteriopathy (GENOA)*

Data management and analysis in GENOA was performed by: Wei Zhao, Jennifer A Smith, Sharon L.R Kardia and Lawerence F Bielak.

*Cameron County Hispanic Cohort (CCHC)*

Miryoung Lee, Joesph B McCormick, Susan P Fisher-Hoch and Kari E North oversaw the consortium and managed the data. Mohanraj Krishnan and Mohammad Yaser Anwar performed the primary genetic analyses in the CCHC study.

*UK BioBank*

Laura M. Raffield and Mohanraj Krishnan performed primary analyses in the UK BioBank.

**PAGE anthropometry working group -** Steven Buyske, Natalie Chami, Stephen S Rich, Kendra R Ferrier, Ethan M Lange, Christopher G Gignoux and Genevieve L Wojcik helped with refinement of study design and plan for analysis. The PAGE Coordinating Center comprises the ARIC, BioMe CARDIA, MEC, HCHS/SOL and WHI studies.

**Advisors**

Kari E North, Marielisa Graff, Ruth J.F Loos, Jennifer A Smith, Leslie A Lange, Mariah C Meyer, Ching-Ti Liu, Lisa R Yanek, Miryoung Lee, Penny Gordon Larsen, Laura M Raffield, Jennifer E Below, Anne E Justice, Roelof A.J Smit and Esteban J Parra oversaw the preparation of the manuscript.
