## Supplementary material for "Genome-wide association study provides novel insight into the genetic architecture of severe obesity": Funding

**Author contributions**

We acknowledge the participants in each cohort contributing to this study. This research was supported by the following funding bodies:

**Stage 1 – Genome‐wide association studies - discovery**

**ARIC** - The Atherosclerosis Risk in Communities Study is carried out as a collaborative study supported by National Heart, Lung, and Blood Institute contracts. The ARIC study has been funded by funds from the National Heart, Lung, and Blood Institute, National Institutes of Health Department of Health and Human Services (contract numbers HHSN268201700001I, HHSN268201700002I, HHSN268201700003I, HHSN268201700004I and HHSN268201700005I), R01HL087641, R01HL086694; National Human Genome Research Institute contract U01HG004402; and National Institutes of Health contract HHSN268200625226C. The following members of the ARIC Research Team contributed to this study: Mohanraj Krishnan, Mohammad Yaser Anwar (supported by R01DK122503 – 02W1), Benjamin S Hadad, Baiyu Qi, Heather M Highland, Kristin L Young, Yujie Wang, Kari E North (R01HL163262) and Mariaelisa Graff (R01HL163262).

**BioMe** – We appreciate all individuals participating in the BioMe TM Biobank Program. This study was led by Michael H Preuss, Ryan W Walker and Ruth J.F Loos (R01DK110113; R01DK075787; R01DK107786; R01HL142302; R01HG010297; R01DK124097; R01HL151152, P30ES023515, P30ESDK020541, R01ES033688, R01DK137968).

**BioVu -** Vanderbilt University Medical Center’s BioVU projects are supported by numerous sources: institutional funding, private agencies, and federal grants. These include the NIH funded Shared Instrumentation Grant S10OD017985 and S10RR025141; CTSA grants UL1TR002243, UL1TR000445, and UL1RR024975. Genomic data have also been supported by investigator-led projects that include U01HG004798, R01NS032830, RC2GM092618, P50GM115305, U01HG006378, U19HL065962, R01HD074711. The following members of the BioVu Research Team contributed to this study: Hung-Hsin Chen, Rashedeh Roshani and Jennifer E Below.

**The Coronary Artery Risk Development in Young Adults Study (CARDIA)** – We greatly appreciate the participants of CARDIA. The Coronary Artery Risk Development in Young Adults Study (CARDIA) is conducted and supported by the National Heart, Lung, and Blood Institute (NHLBI) in collaboration with the University of Alabama at Birmingham (75N92023D00002 & 75N92023D00005), Northwestern University (75N92023D00004), University of Minnesota (75N92023D00006), and Kaiser Foundation Research Institute (75N92023D00003). Genotyping was funded as part of the NHLBI Candidate-gene Association Resource (N01-HC-65226) and the NHGRI Gene Environment Association Studies (GENEVA) (U01-HG004729, U01-HG04424, and U01-HG004446). Myriam Fornage and Penny Gordon-Larsen led the CARIDA component of this study.

**Framingham Heart Study (FHS)** - This research was conducted in part using data and resources from the Framingham Heart Study of the National Heart Lung and Blood Institute of the National Institutes of Health and Boston University School of Medicine. This work was partially supported by the National Heart, Lung and Blood Institute's Framingham Heart Study (Contract Nos. NO1-HC-25195, HHSN268201500001I and 75N92019D00031) and its contract with Affymetrix, Inc for genotyping services (Contract No. N02-HL-6-4278). A portion of this research utilized the Linux Cluster for Genetic Analysis (LinGA-II) funded by the Robert Dawson Evans Endowment of the Department of Medicine at Boston University School of Medicine and Boston Medical Center. We also acknowledge the dedication of the FHS study participants without whom this research would not be possible. This research was partially supported by grant R01-DK122503 from the National Institute of Diabetes and Digestive and Kidney.

**The Hispanic Community Health Study / Study of Latinos (HCHS/SOL)** – We would like to thank all the families who participated in the HCHS/SOL study to identify traits that impact Latino/Hispanic health. HCHS/SOL: Primary funding support to Kari E North and colleagues is provided by U01HG007416. Additional support was provided via R01DK101855 and 15GRNT25880008. The HCHS/SOL study was carried out as a collaborative study supported by contracts from the National Heart, Lung, and Blood Institute (NHLBI) to the University of North Carolina (N01-HC65233), University of Miami (N01-HC65234), Albert Einstein College of Medicine (N01-HC65235), Northwestern University (N01-HC65236), and San Diego State University (N01-HC65237).The following members of the HCHS/SOL Research Team contributed to this study: Mohanraj Krishnan, Mohammad Yaser Anwar (supported by R01DK122503 – 02W1), Benjamin S Hadad, Baiyu Qi, Heather M Highland, Kristin L Young, Yujie Wang, Kari E North (R01HL163262) and Mariaelisa Graff (R01HL163262).

**Mexico City 1 and Mexico City 2 (MC1 & MC2)** - MC1 & MC2 gratefully acknowledge the contributions of the participants. This study cohort data management and initial analysis was led by Esteban J Parra, Miguel Cruz, Frida Lona-Durazo, Jesus Peralta and Jamie Gomez-Zamudio. The Mexico City 1 and Mexico City 2 studies were supported in Mexico by the Fondo Sectorial de Investigación en Salud y Seguridad Social (SSA/IMSS/ISSSTECONACYT, Project 150352), Temas Prioritarios de Salud Instituto Mexicano del Seguro Social (2014-FIS/IMSS/PROT/PRIO/14/34), and the Fundación IMSS. In Canada, this research was enabled in part by two CIHR Operating grants to Esteban Parra, a CIHR New Investigator Award to Esteban Parra and by support provided by Compute Ontario (www.computeontario.ca), and Compute Canada ([www.compute.canada.ca](http://www.compute.canada.ca)).

**Multiethnic Cohort Study (MEC)** - MEC is funded by the National Cancer Institute (U01CA164973, MPI: Loic Le Marchand, Christopher Haiman, Lynne R Wilkens). The cohort was established in 1993-1996 to examine lifestyle risk factors and genetic susceptibility to cancer.

**MyCode/DiscovEHR** - We thank all the participants of the MyCode Study. We thank the members of the Geisinger-Regeneron DiscovEHR Collaboration who have been critical in the generation of the genetic data used in this study. Study authors were all funded by the: R01 DK 122503. The study contributing to data management and GWAS of this cohort includes: Geetha Chittoor, Anne E Justice, Navya S Josyula and Christopher R Bauer.

**Netherlands Epidemiology of Obesity Study (NEO)** – The NEO study was supported by the participating departments, the Division and the Board of Directors of the Leiden University Medical Centre, and by the Leiden University, Research Profile Area ‘Vascular and Regenerative Medicine’. We thank all individuals who participated in the NEO study, and all participating general practitioners for inviting eligible participants. The authors also thank P.R. van Beelen and all research nurses for the collection of data, P.J. Noordijk and her team for sample handling and storage, and I. de Jonge for data management of the NEO study.

**Women’s Health Initiative Study (WHI)** - The WHI program is funded by the National Heart, Lung, and Blood Institute, National Institutes of Health, U.S. Department of Health and Human Services through contracts 75N92021D00001, 75N92021D00002, 75N92021D00003, 75N92021D00004, 75N92021D0000

**Stage 2 – Genome‐wide association studies – replication**

**Genetic Epidemiology Network of Arteriopathy (GENOA)** - We would like to thank the families that participated in the GENOA study. Support for GENOA was provided by the National Heart, Lung and Blood Institute (U01 HL054457, U01 HL054464, U01 HL054481, R01 HL119443, and R01 HL087660) of the National Institutes of Health. Data management and analysis was performed by: Wei Zhao, Jennifer A Smith, Sharon L.R Kardia and Lawerence F Bielak.

**Genetic Studies of Atherosclerosis Risk (GeneSTAR)** – We greatly appreciate all the individuals who participated in the GeneSTAR study. GeneSTAR was supported by grants from the National Institutes of Health/National Heart, Lung, and Blood Institute (U01 HL72518; HL087698; HL49762; HL58625; HL071025); NIH/National Institute of Nursing Research (NR0224103) and by a grant from the NIH/National Center for Research Resources (M01-RR000052) to the Johns Hopkins General Clinical Research Center. All GeneSTAR studies were approved by the Johns Hopkins Medicine Institutional Review Board. Data management of the GeneSTAR program was led by Lisa R Yanek and Dhananjay Vaidya.

**Health and Recruitment Study (HRS)** - HRS is supported by the National Institute on Aging (NIA U01AG009740). The genotyping was funded separately by the National Institute on Aging (RC2 AG036495, RC4 AG039029). Additional funding from US National Institutes of Health grants U01AG009740, RC2 AG036495, RC4 AG039029. This program was coordinated by: Wei Zhao, Jennifer A Smith, Sharon L.R Kardia, Jessica D Faul and David R Weir.

**Million Veterans Program (MVP)** - VA Million Veteran Program (MVP) The MVP gratefully acknowledges the contributions of the participants and of the study staff. This research is based on data from the Million Veteran Program, Office of Research and Development, Veterans Health Administration, and was supported by awards: I01-BX004821 and I01-BX005831. This publication does not represent the views of the Department of Veteran Affairs or the United States Government.

**REasons for Geographic and Racial Differences in Stroke (REGARDS) –** This research project is supported by cooperative agreement U01 NS041588 co-funded by the National Institute of Neurological Disorders and Stroke (NINDS) and the National Institute on Aging (NIA), National Institutes of Health, Department of Health and Human Service. The content is solely the responsibility of the authors and does not necessarily represent the official views of the NINDS or the NIA. Representatives of the NINDS were involved in the review of the manuscript but were not directly involved in the collection, management, analysis or interpretation of the data. The authors thank the other investigators, the staff, and the participants of the REGARDS study for their valuable contributions. A full list of participating REGARDS investigators and institutions can be found at: <https://www.uab.edu/soph/regardsstudy/>.

**UK Biobank (UKBB)** - We are grateful to the UK Biobank participants. This research has been conducted using the UK Biobank Resource under project 12505.
Laura M Raffield was funded in part by NIH KL2TR002490.

**Other studies contribution to the genetic analysis of severe obesity.**

**PAGE** – We would like to thank Steven Buyske being involved in the PAGE Coordinating Center which comprises the ARIC, BioMe CARDIA, MEC, HCHS/SOL and WHI studies.

**Cameron County Hispanic Cohort (CCHC)** - We thank the participants from the CCHC study. The authors would like to thank the CCHC cohort team, Joesph B McCormick and Susan P Fisher-Hoch who contribute their insights into this study. This work was funded in part by AHA grant 903805, NIH awards: NIH/NHLBI R01HL142302, R01HL151152, R01 DK122503, R01HD057194, R01HG010297, R01HL143885, NIH/NIDDK R01DK127084, R21DK122234. This study was also funded in part by the Center for Clinical and Translational Sciences UM1TR004906) from the National Center for advancing Translational Sciences, National Institutes of Health Clinical and Translational Award grant no. UL1 TR000371 from the National Center for Advancing Translational Sciences, Multi-Omics for obesity-associated liver discovery in Hispanics/Latinos: the Cameron County Hispanic Cohort (U01CA288325) and Integrative multi-omics for discovery of molecular pathways associated with diabetic retinopathy (R01EY036258).
