## Supplementary material for "Genome-wide association study provides novel insight into the genetic architecture of severe obesity": Studies

**Study populations.**

A. **Discovery studies**

**The Population Architecture using Genomics and Epidemiology (PAGE) Consortium**

PAGE^1^ is a collection of studies across multiple ethnicities and backgrounds funded by the National Institutes of Health to examine the genetic architecture of common complex diseases and traits in diverse populations. PAGE data used in this analysis included participants enrolled in the following cohort studies:

*Atherosclerosis Risk in Communities Study (ARIC)*

ARIC is a multi-center prospective epidemiologic cohort study funded by the National Heart, Lung, and Blood Institute (NHLBI) to investigate the causes of atherosclerosis and its clinical outcomes, and variation in cardiovascular risk factors, medical care, and disease by race, gender, location, and date^2^. In total, 15,792 participants ages 45-64 of primarily European American and African American descent were recruited between 1987 and 1989 from four communities in the United States: Washington County, MD; Forsyth County, NC; Jackson, MS; and Minneapolis, MN. At study baseline (1987-1989), participants received standardized physical examinations and interviewer-administered questionnaires. Semi-annual telephone follow-up calls are ongoing to maintain contact and assess the health status of the cohort.

*The BioMe^TM^ Biobank Program (BioMe)*

Funded by the Charles Bronfman Institute for Personalized Medicine at Mount Sinai Medical Center (MSMC), BioMe is an EMR-linked biorepository drawing from Mount Sinai Medical Center consented patients which were drawn from a population of over 70,000 inpatients and 800,00 outpatients annually from diverse local communities in upper Manhattan (https://icahn.mssm.edu/research/ipm/programs/biome-biobank). Data on anthropometrics, demographics, and medication use were derived from participants’ EMR and a medical history questionnaire administered at baseline^3^.

*The Coronary Artery Risk Development in Young Adults Study (CARDIA)*

CARDIA is a multi-center prospective cohort study funded by the NHLBI to study the development and distribution of cardiovascular diseases and their risk factors^4^. A total of 5,115 participants ages 18-30 years (52% African American, 55% women) were recruited in 1985-1986 from four communities in the United States: Birmingham, AL; Chicago, IL; Minneapolis, MN; and Oakland, CA. The participants were selected so that there would be approximately the same number of people in subgroups of race, gender, education (high school or less/more than high school) and age (18-24 and 25-30) in each of these 4 centers. Participants were asked to participate in follow-up examinations during 1987-1988 (Year 2), 1990-1991 (Year 5), 1992-1993 (Year 7), 1995-1996 (Year 10), 2000-2001 (Year 15), and 2005-2006 (Year 20), 2010-2011 (Year 25), 2015-2016 (Year 30), and 2020-2022 (Year 35). Data have been collected on factors believed to be related to heart disease, including blood pressure, cholesterol and other lipids, and glucose as well as physical measurements such as weight and skinfold fat, as well as lifestyle factors, behavioral and psychological variables, and medical and family history.

*The Multiethnic Cohort Study (MEC)*

MEC is a prospective cohort study funded by the National Cancer Institute (NCI) to study diet and cancer in the United States^5^. In total, 215,251 participants living in Hawaii and California (primarily Los Angeles County) ages 45-75 years were recruited between 1993 and 1996 (16.3% African American, 22.0% Latino, 26.4% Japanese American, 6.5% Native Hawaiian, 22.9% white). Upon recruitment, participants completed a self-administered questionnaire on demographic, dietary, and lifestyle traits. Biological specimens were also 3 collected from over 70,000 MEC members in 2001-2005. There were seven ancillary MEC sub-studies included in this analysis: the Slim Initiative in Genomic Medicine for the Americas (MEC-Sigma), a type 2 diabetes study in Hispanic/Latino adults; MEC-AAPC, -JAPC, and - LAPC, studies of prostate cancer in African American, Asian, and Hispanic/Latino men, respectively; and MEC-AABC, -JABC, and LABC, studies of breast cancer in African American, Asian, and Hispanic/Latina women, respectively.

*The Hispanic Community Health Study/Study of Latinos (HCHS/SOL)*

HCHS/SOL is a multi-center, community-based cohort study of Hispanic/Latinos in the United States to identify the prevalence of and risk factors for multiple chronic diseases, including cardiovascular diseases, lung, kidney and liver diseases^6^. Over 16,000 participants ages 18-74 years were recruited from four communities in the United States between 2008 and 2011: Bronx, NY; Chicago, IL; Miami, FL; and San Diego, CA. These recruitment sites were selected so that the overall sample would include at least 2,000 people from each of the following origin designations: Mexican, Puerto Rican and Dominican, Cuban, and Central and South American. Households were selected via a two-stage sampling within census block groups^7^. At baseline (2008-2011), participants received standardized examinations and interviewer administered questionnaires. A re-examination of the HCHS/SOL cohort was conducted during 2015-2017, and annual telephone follow-up interviews are ongoing since study inception to determine health outcomes of interest.

*The Women’s Health Initiative Study (WHI)*

WHI is a long-term prospective study to investigate causes of morbidity and mortality among postmenopausal women in the United States^8^. Between 1993 and 1998, 161,808 4 women ages 50-79 years were recruited from 40 clinical centers and enrolled in randomized clinical trials or an observational cohort study. Women in the observational study received a standardized examination at baseline and interviewer-administered questionnaires. The following ancillary studies were included in this analysis: the Genetics and Epidemiology of Colorectal Cancer Consortium (GECCO), the Modification of PM-Mediate Arrhythmogenesis in Populations study (MOPMAP), the Genomics and Randomized Trials Networks (GARNET), the Hip Fracture GWAS (HIPFX), the Long-Life Study (LLS), the Women’s Health Initiative Memory Study (WHIMS), and the Women’s Health Initiative-SNP Health Association Resource (WHI-SHARe). The GECCO study aims to investigate the genetic susceptibility of colorectal cancer, including common and rare loci, gene-environment interactions, and survival. The MOPMAP study aims to investigate the susceptibility to arrhythmogenic effects of particulate matter air pollution contributed by common genetic and environmental variation. The GARNET study is a series of genome-wide association studies of treatment response in randomized clinical trials, aiming to identify genetic variants associated with response to treatments for conditions of clinical or public health significance. The HIPX study was designed to perform epidemiological studies of hip fracture in women. The LLS study included 7,875 women from the WHI Extension II Medical Records Cohort (MRC). The LLS consisted of a one-time in-person visit (between March 2012 and May 2013) with a blood draw, a brief clinical assessment, and an assessment of functional status. The WHIMS study is a trial to examine the effect of oestrogen therapy in preventing and slowing the progression of dementia. The WHI-SHARe study is part of NHLBI’s SNP Health Association Resource (SHARe) project, aiming to enhance the statistical power for research specific to groups defined by race and ethnicity and to discover or replicate genes associated with quantitative traits (such as blood pressure and blood lipids) in 5 these groups. The participants in GECCO, MOPMAP, GARNET, HIPFX, LLS and WHIMS are self-reported European Americans, while the participants in WHI-SHARe are self-reported African Americans.

**Mexico City Studies (MC1 & MC2)**

The MC1 and MC2 studies were designed as case-control studies of type 2 diabetes (T2D). Both studies sampled individuals from Mexico City who were covered by the Mexican Social Security Institute (IMSS). Extensive phenotype data was available for these samples, including T2D status, age, sex, BMI, WHR, lipid levels and blood pressure, among others. The two samples were described in Below et al. 2016 ^9^.

**Framingham Heart Study (FHS)**

In 1948, researchers recruited 5,209 men and women between the ages of 30 and 62 from Framingham, MA, as the original cohort. Participants then examined approximately every 2 since 1948 in order to obtain updated information on potential CVD risk factors and lifestyle habits ^10^. In 1971, the study enrolled a second-generation cohort -- 5,124 of the original participants' adult children and their spouses -- to participate in similar examinations. This “offspring cohort” was then examined approximately every 4-8 years since 1971. In April 2002, the enrollment of a third generation of participants, the grandchildren of the original cohort. It involved 4,095 participants, as Gen3. This Gen3 cohort was examined approximately every 4-8 years.

**Netherlands Epidemiology of Obesity (NEO)**

The Netherlands Epidemiology of Obesity (NEO) is a population-based study consisting of 6,671 individuals living in the greater area of Leiden, the Netherlands, aged between 45 to 65 years ^11^. This study is designed to investigate mechanisms that lead to common complex disease and thus has an oversampling of individuals with a BMI greater than 27kg/m^2^. Participants eligible for this study were asked to collect their urine over a 24-hour period and complete a general questionnaire to report demographic, lifestyle and clinical information. At baseline visit, other physical characteristics were collected including anthropometric measurements, blood pressure, both fasting and postprandial blood sampling (30 minutes and 2.5 hours after a liquid mixed meal), ECG, carotid artery IMT, and pulmonary function tests.

**BioVU**

The main objective of the BioVU project was to develop a DNA biobank to link phenotypic data derived from the electronic medical record (EMR) system led by Vanderbilt University, Tennessee^12^. Surveys were sent to potential participants and approximately 90% of respondents were comfortable with the concept of anonymized genetic data being used for research. By the end of 2010, approximately 130,000 samples were collected and de-identified. The most common diagnoses were hypertension (15.7%), type II diabetes (11.8%), hyperlipidemia (11.5%), coronary artery disease (7.8%), and anemia (5.9%). Most (88%) records have at least one medication indicated, with an average of 8.0 ± 6.8 medications per record. DNA was extracted from surplus blood samples and genotyping data was linked to de-identified EHR data.

**MyCode/DiscovEHR**

The MyCode™Community Health Initiative (MyCode) study is a healthcare-based population study in central and northeastern Pennsylvania with ~2 million patients^13^. All participants provided informed consent for participation in the MyCode Study. This study was approved by the Geisinger Institutional Review Board. The DiscovEHR study includes a subset of the MyCode participants that have been genotyped to enable the use of EHR data for discovery research^14^. For this study, we used data from self-identified Europeans, Africans, Asians, and Hispanic/Latino participants. At the time of analysis, DiscovEHR included 92,476 consented individuals with genetic data. Genotyping and quality control of DiscovEHR data have been previously described^15^. Briefly, of the 92,476 with array-based genotyping, 67% were genotyped using the Illumina HumanOmniExpressExome (HOEE) genotyping platform with remainder typed on the Illumina Global Screening Array (GSA). These data are processed with Illumina’s GenomeStudio, imputed to the 1000 Genome Phase III. All array-based data are QC’d with standard quality control procedures before association testing.

**B. Replication or Follow-up analyses**

**Million Veteran Program (MVP)**

The Million Veteran Program (MVP) is a national research program to learn how genes, lifestyle, and military exposures affect health and illness. Between 2011 and January 2023 more than 900,000 U.S. Veterans (90% men, 10% women) have joined one of the world's largest programs on genetics and health^16^. Approximately 70% of the participants are White, 20 percent are Black, and 8% are Hispanic. Data resources include linkage to the Veterans electronic health record, genetic information based on a 750,000-SNP Biobank chip, questionnaires administered at the time of recruitment, and linkage of outcomes to Medicare/Medicaid and the National Death Index.

**REasons for Geographic and Racial Differences in Stroke (REGARDS)**

The Reasons for Geographic and Racial Differences in Stroke (REGARDS) project, sponsored by the National Institutes of Health (NIH), is a national study focusing on learning more about the factors that increase a person's risk of having a stroke^17^.

REGARDS is an observational study of risk factors for stroke in adults 45 years or older. 30,239 participants were recruited between January 2003 and October 2007. They completed a telephone interview followed by an in-home physical exam. Measurements included traditional risk factors such as blood pressure and cholesterol levels and an electrocardiogram of the heart. At six-month intervals, participants are contacted by phone to ask about stroke symptoms, hospitalizations, and general health status. The study is ongoing and will follow participants for many years."

**Genetic Studies of Atherosclerosis Risk (GeneSTAR)**

GeneSTAR is a prospective study which was designed to examine environmental, phenotypic, and genetic causes of premature cardiovascular disease. Participants came from European- and African American families identified from probands with a premature coronary artery disease (CAD) event prior to 60 years of age who were identified at the time of hospitalization in any of 10 Baltimore area hospitals from 1983-2006^18^. Their apparently healthy 30–59-year-old siblings without known CAD were recruited and underwent phenotypic measurement and characterization between 1983 and 2007. From 2003-2006, adult (age ≥ 21) offspring of all participating siblings and probands along with the coparent of the offspring were recruited and underwent phenotypic measurement and characterization. Weight was measured in pounds using a clinical balance scale with participants wearing light, indoor clothing. Height was measured in inches using a stadiometer. Data from the first visit was used for all analyses.

**Health and Retirement Study (HRS)**

The University of Michigan Health and retirement study (HRS) is a nationally representative longitudinal panel study that surveys more than 37,000 individuals over age 50 in 23,000 households in the USA ^19^. An overrepresentation of Blacks and Hispanic individuals, particularly Mexican Americans and who reside in Florida were recruited to participate in this study. Information obtained in 2006 face to face interviews included physical health characteristics (interviewer-measured height, weight, waist circumference, and blood pressure), physical performance measures, and biomarkers (e.g., blood spots to test for cholesterol, C-reactive protein, and hemoglobin A1C). Saliva samples were collected as the main source of genetic material, and approximately 2.4 million variants were genotyped in 15,000 respondents in either 2011, 2012 or 2015.

**Genetic Epidemiology Network of Arteriopathy (GENOA)**

GENOA is one of four research networks that form the NHLBI Family Blood Pressure Program (FBPP)^20^. From its inception in 1995, GENOA's long-term objective was to elucidate the genetics of hypertension and its arteriosclerotic target-organ damage, including both atherosclerotic (macrovascular) and arteriolosclerotic (microvascular) complications involving the heart, brain, kidneys, and peripheral arteries. Two GENOA cohorts were originally ascertained (1995-2000) through sibships in which at least 2 siblings had essential hypertension diagnosed prior to age 60 years. All siblings in the sibship were invited to participate, both normotensive and hypertensive. These include non-Hispanic White Americans from Rochester, MN (n =1583 at the 1st exam) and African Americans from Jackson, MS (N=1854 at the 1st exam). During the second exam (2000-2005), approximately 80% of participants were re-recruited. The GENOA data consists of biological samples (DNA, serum, urine) as well as demographic, anthropometric, environmental, clinical, biochemical, physiological, and genetic data for understanding the genetic predictors of diseases of the heart, brain, kidney, and peripheral arteries.

**UK Biobank (UKBB)**

The UK Biobank (UKBB) is a large population based prospective biomedical study comprising in depth genetic determinants and health information from approximately 500,000 individuals recruited in 22 assessment centers across England, Wales and Scotland between 2006 and 2010 ^21^. Genome-wide genotyping was performed on all UK Biobank participants using the UK Biobank Axiom Array. Approximately 850,000 variants were directly measured, with > 90million variants imputed using the Haplotype Reference Consortium and UK10K + 1000 Genomes reference panels.

**Cameron County Hispanic Cohort (CCHC)**

The CCHC is a randomly ascertained, community-based cohort of Mexican Americans recruited from border communities on the Texas-Mexico border^22^. stablished in 2004, this large cohort study, currently numbering around 5,000 individuals, documents sociodemographic, clinical, behavioural, and biologic characteristics of Cameron County Mexican Americans. Households were randomly identified by US census tract/block and members were invited to participate in the study. The baseline examination included physical examination, collection of biospecimens, and completion of questionnaires.
